# Cardiac Implantable Electronic Devices’ longevity: A novel modelling tool for estimation and comparison

**DOI:** 10.1101/2025.03.30.25324901

**Authors:** Pascal Defaye, Serge Boveda, Jean-Renaud Billuart, Klaus K Witte, Maria F Paton

## Abstract

**Aims:** Generator longevity is the key issue for patients, and is also important for payers, yet implanters of Cardiac Implantable Electronic Devices (CIEDs) face a challenge when selecting the appropriate device since battery longevity is only known for previous generation devices and whilst projected longevities are available for current devices, these are not in comparable formats. This study presents a new framework that facilitates an estimation of longevities for all CIEDs of both previous and existing generations that could simplify personalization of the device choice.

**Methods:** Longevity can be calculated based upon a simple concept entitled the “power consumption index” (PCI = t x I/C, where t is a constant of 1 hour, I is the current required by the device and C, its battery capacity). We retrieved published data from the user manuals of all commonly used pacemakers including single chamber, dual chamber, cardiac resynchronization and leadless devices. C and the components of current I including background current (I_background_) and the pacing current (I_pacing_) were calculated prior to calculation of the PCI for each device. Subsequently, a set of fictitious patient pool conditions via a Monte-Carlo simulation were used to model CIED survival curves which were then compared with real-life data from the Swedish device registry of previous generation CIEDs. Finally, we modeled survival curves for current generation devices using the PCI model.

**Results:** Using the PCI approach we were able to calculate longevities for all pacemaker devices under a variety of settings. The modeled I_background_ matched the data reported by manufacturers, and, under a variety of settings, regression analysis showed a low average error rate between industry-reported and modelled longevities (ratio: modelled longevity/industry reported longevity -1) = 0.1±4.0% and 0.1±0.7% for previous and existing SR/DR devices, 1.0±5.0% and 0±3.0% for previous and existing CRT-P, and 0±4.0% for leadless pacemakers, respectively). More than 50% of the PCI and thereby a significant contributor to longevity was accounted for by I_background_. I_pacing_ was the second largest contributor (20% for standard single and dual chamber devices, 30% for CRT-P and 40% for leadless devices). Certain pacing algorithms and IEGM storage considerably impacted specific devices with longevity losses of up to 1 year. The Monte-Carlo analysis demonstrated consistency between projected longevities by the PCI model and real-life data for historical devices and the calculated longevities that stemmed from this were consistent with the real-world data from Sweden.

**Conclusion:** The PCI model combining power consumption and battery capacity allows a comparison of longevity across CIEDs and programming options. Such a tool could help implanters improve personalization of device prescription for their patients and payers to make more informed decisions about tailoring device purchases and programming most appropriate for their population.

**What’s new?:** The Power Consumption Index (PCI) is a new approach to estimating the longevities of CIEDs across models, manufacturers, settings and pacing options that allows comparisons across devices and manufacturers.

## Introduction

Longevity of Cardiac Implantable Electronic Devices (CIEDs) is an important issue for patients, who wish to avoid further surgery, and purchasers, who wish to optimize cost-effectiveness, and is therefore a relevant consideration for clinicians. It is appreciated that there are common discrepancies between declared (future) longevities of generators and their subsequent survival curve once implanted (1,**Error! Reference source not found.**). Despite calls for more transparency and industry-wide standardized reporting of longevity (3,4,5), comparisons of longevity between devices and manufacturers in different settings remains challenging. Given that 30-40% of all CIED procedures are generator replacements, there exists the risk of conflicts of interest for both manufacturers, and, in fee-for-service healthcare environments providers, that limit enthusiasm for transparency (6).

Although implanters and their patients appreciate the concept of battery capacity as a prime criteria for device longevity (7) they also recognize that energy drain plays a role (8,9). However, the potential lifetime of the device is also determined by how energy is stored, and how efficiently it is delivered(10), along with the patient’s characteristics, all of which, add to the frustrating situation of complex and non-standardized user-manual declared longevity, with different programming as baseline across companies, making personalization of generator prescription impossible even in the presence of similar battery capacity. If one could reliably describe consumption and index this to battery capacity and pacing requirements, there remains the possibility of a reliable comparison of devices.

Based upon previous work defining the power consumption of CIEDs,(11) which included a calculation for the inverse of longevity, we have developed the Power Consumption Index (PCI) (as defined by PCI = t x I/C (where t is a constant equal to 1 hour)) that aims to describe the intrinsic power consumption of the overall system (the pacing system coupled with the battery) during a normalized period (1 hour). The reciprocal of the PCI therefore allows a derivation of longevity (Figure 1).

Hence, the objectives of this study were to create and test a “universal” model, based on the concept of PCI, that offers the opportunity to model longevity for any CIED. The model was subsequently validated by comparing the modeled survival curves of previous generation CIEDs with real life data and then applied to predict product survival curves for new generations of CIEDs.

## Methods

Firstly, we collected battery capacity and estimated current drain for a variety of devices from manufacturer’s user manuals to calculate PCI values and thereby longevity. The user manuals are available from the five major CIEDs’ manufacturers: Abbott (Abt, formerly St Jude Medical, Sylmar, CA, USA), Boston Scientific (BSc, St Paul, MN, USA), Biotronik (Btk, Berlin, Germany), Microport CRM (Mcp, formerly LivaNova and Sorin, Clamart, France), and Medtronic (Mdt, Minneapolis, MN, USA). We also used the only web-based longevity calculator available from BsC (12). Detailed results are provided in *Supplementary Materials 1*.

### Battery capacity

Nominal voltage and battery capacities at ERI were systematically retrieved from user manuals for single and dual-chamber (SR and DR) pacemaker, cardiac resynchronization therapy (cardiac resynchronization therapy with pacemaker [CRT-P]) and leadless pacemaker (PM) models. Representative models of previous and new generations of CIEDs were selected based on experts’ opinion (*Table 1: Devices models)*.

### Current drain modelling

Industry-reported CIEDs’ longevities in user manuals depend upon the programming (including the activation of specific algorithms such as rhythm storage, remote monitoring, and sensors). Current is not provided in manuals and therefore modeling was required to calculate the PCI for each CIED. Because nominal settings differ from one manufacturer to another, an additional step was necessary. This involved identifying and estimating the current for background and pacing (I_background_, I_pacing_) and that required for the optional settings (I_remote_ I_IEGM_ I_algo_). For I_background_, and I_pacing_ the evaluation was done by a regression analysis. For I_remote/IEGM/algo_ the current was estimated via the difference of longevity between activated and deactivated settings for each option.

Two categories of optional settings were considered. The first considered algorithms directly influencing the I_pacing_ drain such as automatic threshold management or reduction of right ventricular pacing (RVP) percentage (I_algo_) and the second explored optional settings such as remote monitoring and IEGM storage (I_remote_ and I_IEGM_). Unlike I_background_ or basic pacing, each factor needed to be analyzed individually (*Supplementary Materials 1)*.

Different pacing options were considered. For 20 previous generation single chamber and dual chamber pacemakers, 674 settings were considered, for 11 new generation devices 243 settings were explored and for 8 previous and 5 current CRT-P devices, respectively 294 and 177 settings and for 3 leadless devices 156 settings were considered.

### Estimation of previous devices’ nominal longevities, using the PCI model

A literature review was performed to retrieve standard pacemaker parameters (for example, heart rate, threshold, pulse amplitude, pulse duration) in clinical studies, registries and clinical practice. Data related to specific algorithms (reduction of ventricular pacing (hysteresis and MVP^TM^, aCRT^TM^), automatic management of pacing output) were also collected via clinical studies, when available. Where the effect on RV pacing of the algorithm was missing, for example with hysteresis and adaptive CRT, these were (imputed) using data provided for clinical practice and assumed to be stable over the lifetime of the device. We also assumed that managed output algorithms would achieve and maintain the ideal of 1V (albeit at cost as described below). Then, under nominal conditions, the PCIs and the corresponding nominal longevities were estimated for the CIEDs listed below (details in *Supplementary materials 1*) via the battery capacity values and the current drain modeling (via the formulas **PCI = t x I/C** and **L = 1/PCI)**

### Validation of PCI model, using Monte-Carlo simulations

In order to validate the survival curves the model was applied not only according to nominal parameters but a variety of settings due to reflect real-life patient situation and programming. A pool of fictitious patients’ sets (100,000 patients) was created via a Monte-Carlo simulation (programmed in Python™) with age, indication (sinus node dysfunction [SND], intermittent atrio-ventricular block [AVB], complete AVB) and programmed parameters based on available literature (14,15,16,17,18,19,20,21). The parameters used for the Monte-Carlo model are described in *Supplementary Materials 2*. Right ventricular pacing avoidance [RVPa] algorithms were assumed to be applied for SND patients eligible for DDD pacing. Remote monitoring was not standard for previous generation devices. We hypothesized a 50% adoption rate of remote for new generation current devices. The impact of settings (such as capture management, additional IEGM storage, RVPa for intermittent AVB for pacemaker, aCRT™ and MPP™ for CRT) on energy consumptions were studied.

For each device, longevity was calculated per patient using via the PCI energy consumption formula. Missing information which could not be derived from manuals was hypothesized, while assuming similarities among same generation devices. When longevity exceeded patient life expectancy, data were censored (since end of service uncommonly occurs simultaneously and residual battery life of the device is rarely collected at death). The distribution of PCI and corresponding longevity across the pool of fictitious patients allowed the drafting of product survival curves for each cardiac implant. The PCI model was then validated for previous generation’s devices by comparing these modeled product survival curves and real-life data. For the development of the real-world device ‘survival’ curve, we used the Swedish Pacemaker and ICD registry [13] which was started in 1989 on the initiative of the Swedish Society of Cardiology. All the implanting clinics in Sweden report to this registry which compiles quarterly and annual reports of pacemaker use in Sweden. Each year they register about 5000 pacemaker procedures. The real-world product survival curves were extracted from these open-access annual reports.

### Estimation of current devices’ longevity, using the PCI model

Finally, the PCI model was used to forecast survival curves for contemporary devices, using the same method as the one described previously.

## Results

### Battery capacity retrieved from manufacturers’ manuals

Battery capacities for single and dual chamber pacemakers have remained, on average, unchanged between previous generation and contemporary devices (close to 1 Ah), with disparities among manufacturers (Table 2).

In CRT-P devices, most manufacturers use the same battery capacity as for their conventional pacemaker, close to 1 Ah. Only BSc offers a battery above 1 Ah (using the same technology as Accolade™ DR EL) with 1.5 Ah at elective replacement indicator (ERI).

Battery capacity for single and dual chamber leadless pacemakers is very different from standard pacemakers. For VVI pacing, the Micra™ capsule (Mdt) is equipped with a 0.12 Ah battery, while the Aveir™ capsule (Abt) battery reached 0.24 Ah. Aveir™ is also available in a dual chamber configuration with the same ventricular capsule combined with a smaller atrial capsule (equipped with a 0.17 Ah battery capacity).

### Current drain modeling

#### Background current (I_background_) and pacing current (I_pacing_)

For conventional pacemakers, the difference between industry-reported longevities and those derived by regression reached 0.1 ±4% for previous generation devices and -0.1 ±0.7% for new generation devices. For devices with a variety of configurations (Mdt and BSc via its longevity calculator website), the regression coefficient (R^2^) exceeded 90%, across all settings applied.

The I_background_ derived by regression analysis (Table 3) matched those reported by manufacturers for most devices with few exceptions. There was, however, a significant change between previous and new generation devices. Previous generation devices relied on a <u>I_background_</u> exceeding 9-10 µA (except for Identity™ for which the <u>I_background_</u> ranged between 5.72 µA and 6.19 µA, the Evia™-T for which the <u>I_background_</u> ranged between 6 µA and 6.66 µA and Symphony™ with an <u>I_background_</u> at 6µA). For new generation CIEDs, the <u>I_background_</u> did not exceed 7 to 7.5 µA, except for BSc devices, which reached 9.7 µA to 10.4 µA. Similar results were observed for CRT-P. For leadless pacemakers, the <u>I_background_</u> was much lower than for conventional pacemakers (ranging from 0.8 µA to 0.94 µA for VVI and 1.75 µA for DDD, including 0.81 µA to 1µA for communication between the atrial and the ventricular capsules).

The analysis conducted in *Supplementary Materials 1 (Pacing current)* shows that for the following settings (60 bpm, pulse 0.4 ms, 500 ohms, 100% pacing), the I_pacing_ was relatively consistent between all categories of contemporary pacemakers with average current drains of 1.98 ±0.12µA at 2,5V pacing outputs and 4.37 ±0.24 µA, at 3.5V. This analysis can also be considered as a sensitivity analysis regarding pacing current parameters.

#### Current from optional settings (I_algo/remote/sensorIEGM_)

Reduction of ventricular pacing algorithms (RVPa) or Adaptive CRT (aCRT^TM^) directly modulate the percentage of RV pacing. Depending on the percentage of reduction of pacing achieved, I_pacing_ decreases from 1.98 - 4.37 µA, respectively at 2,5V and 3,5V (100% pacing), down to 0,89-1,97µA (45% of pacing) and 0,59-1,31µA (30% of pacing). User manuals report that these savings are achieved without energy cost.

On the other hand, multipoint point pacing for CRT (MPP^TM^) increases I_pacing_ on the left ventricular channel up to 3,96 - 8,74µA respectively at both 2,5V and 3,5V.

Threshold algorithms whose sole objective is to guarantee capture, increase pacing outputs and pacing current (user manuals do not specify impact on longevity). Other automatic threshold algorithms aim at optimizing outputs and do this either daily (Capture Management^TM^ from Mdt and Capture Control^TM^ from Btk) or on beat-to-beat basis (Auto capture^TM^ from Abt, Automatic capture^TM^ from Bsc). User manuals do not report an energy cost for the daily algorithms but an energy cost of 1 µA can be derived for beat-to-beat algorithm. The analysis conducted for BsC device (see *Supplementary Materials 1 Algorithms influencing pacing current)* shows that beat-to-beat algorithm saves energy (current drain) only if percentage of pacing is high (>60%) or outputs exceed 2,5V without the algorithm turned on.

Rate adaptive pacing usually relies on a G-sensor (accelerometer) to adapt pacing rate according to effort. Adaption of pacing rate can be optionally enhanced with the combination of a Minute Ventilation (MV) sensor. User manuals describe an estimation of the energy consumption by the MV sensor of (0.69 µA-0.77 µA). The impact of rate adaptive technology on pacing is unknown and somewhat unpredictable.

For most suppliers, IEGM storage is embedded as a standard function and the energy cost related to EGM is included already in the current background. Only Mdt reports a specific impact on longevity (*see Supplementary Materials 1 IEGM*). While IEGM 6-month storage remains minimal (0,11 – 0,34 µA for Enpulse^TM^, 0,04 – 0,11 µA for Azure^TM^), the optional additional use of pre-arrhythmia EGM storage increases current drain and reduces projected service life by approximately by 34% or 4 months per year for Enpulse (equivalent to 5,67-6,16 µA for Enpulse^TM^ 1,3-1,7µA for Azure^TM^).

For remote monitoring, for 2-4 transmission per year, the current consumption is around 1,14-1,75 µA for RF solutions (BsC, Btk) while it is 0,09-0,59 µA for Bluetooth connectivity (Mdt, McP). Btk provides a unique solution as its devices transmit data daily (alerts are managed via its website) with a fixed energy cost close to 1,75 µA.

### Estimation of nominal longevities, using the PCI model

After deriving current drain, the PCIs were computed and corresponding longevities were modelled, at nominal settings, among all devices. Standard settings, PCI value per device and corresponding longevity, evolution between previous and new generation devices are reported in *Supplementary Materials 1*. Figure 2 compares each device per category (VVI, DDD(R), CRT-P) according to the PCI chart presented previously.

Across all devices, the PCI and the corresponding estimated nominal longevity values range from 26.9 (4.2 years) (Enpulse) and 7.6 (15.1 years) (Aveir V with an output of 1.5V).

On average, the PCI for conventional pacemakers reduced between previous and current generations leading to an increase in longevity by more than one year for both SR and DR (10.8 years *vs*. 15.4 years for SR and 11.2 years *vs*. 14.3 years for DR). Unlike SR or DR, on average PCI for CRT-P increased (from 12,5 to 14,1) with the introduction of remote monitoring leading to a reduction of longevity (from 8,3 years to 7,8 years). For leadless devices, the PCI reached 14.2 (corresponding to a longevity of 8.8 years) for dual chamber and 11.7 (10.6 years) for single chamber showing the consequence of energy cost of transmission between capsules in the two-chamber system.

The split of PCI per current highlights a strong impact of the <u>I_background_</u> for all categories of pacemaker: more than 50% of PCI is due to <u>I_background_</u>. The reduction of total PCI for SR/DR between previous and new generations of conventional CIEDs resulted primarily from the reduction of the <u>I_background_</u>. For conventional pacemakers, I_pacing_ accounted for 20% of the total PCI for SR/DR pacemakers and for 30% of the PCI for CRT-P.

Among the contemporary devices, Accolade™ SR/DR had the highest PCI, and thus the lowest estimated longevity (PCI of 13.5 and longevity of 8.5 years for SR, PCI of 14.2 and longevity of 8 years for DR). This is because this device had the highest <u>I_background_</u> (10µA) as compared with other devices of its category. On the other hand, Accolade™ DR EL, even with remote monitoring turned “On”, had the lowest PCI (8.9), and thus the longest longevity (12.8 years), thanks to the high battery capacity at 1.6Ah.

For the other devices, differences were primarily due to optional settings such as extended IEGM, sensor or remote monitoring. In the past, IEGM storage negatively impacted Mdt device longevity. This has been significantly improved upon for both SR and DR devices. Moreover, remote monitoring power consumption is three times higher for RF solutions than Bluetooth solutions (PCI for remote monitoring 1.2 vs 0.4). For example, the estimated longevity for Edora™ 8 reached 9.7 years for SR and 9.1 years for DR. Azure™ benefits from a low remote monitoring power consumption from Buetooth and reached 12.7 years for SR and 11.7 years for DR (extended IEGM turned “Off”). Interestingly, Alizea™ SR and DR benefit from additional battery capacity if remote monitoring is switched off, such that nominal longevity reached 12.9 years, similar to that of the Accolade™ DR EL device. The impact of the sensor was not different between devices.

For leadless SR and DR devices, assumptions included a basic rate of 60bpm, a pulse duration of 0,25 ms for Micra™ and 0,4 ms for Aveir™, an impedance of ∼600 ohms for ventricular pacing and ∼300 ohms for atrial pacing (22,23,24,**Error! Reference source not found.**). Pacing outputs were not reported in studies and two options were considered: 1.5V or 2.5V, reflecting the level of confidence of practitioners in adapting output (thresholds observed were typically low: 1.25 V at implant and 0.75 V weeks after). Pacing percentages were the same as the one for conventional pacemakers. Hysteresis mode was applied for DDD.

Unlike conventional pacemakers, PCI related to pacing accounted for more in the total PCI (40% on average and higher with 2.5V outputs). This is the consequence of a lower battery capacity and thus, a greater proportion of total available energy is required for pacing. Consequently, PCI and longevity significantly changed depending on pacing output assumptions (Aveir™ VVI: 15.1 years for 1.5V *vs*. 10.6 years for 2.5V; Micra™ VVI:10.2 years *vs*. 6.6 years, respectively; Aveir™ V DDD: 12.5 years *vs*. 10.6 years; Aveir™ A DDD 7.3 *vs*. 5.4 years, respectively).

For CRT-P, biventricular pacing is the standard pacing mode with alternative options such as aCRT^TM^ or MPP^TM^ pacing. Biventricular pacing with the Visionist CRT-P device provides a PCI 10.7 and a longevity of 10.6 years, whereas the Percepta™ device with aCRT activated achieves a PCI of 12.3 and longevity of 9.3 years. Not surprisingly Quadra Allure™ with MPP™ activated suffered a considerable increase in PCI and a corresponding reduction in longevity (PCI: 16.9; longevity of 6.8 years).

### Validation of PCI model, using Monte-Carlo simulations

#### Previous generation devices

Modeled survival curves with standard assumptions fitted Swedish registry data for multiple models with few exceptions *(Supplementary Materials 2)*. For conventional pacemakers, modeled survival curves departed from real-life data for following models: Enrythm™ DR, Evia™ DR-T and Identity™ DR 5370. For Enrythm™, the programming of IEGM storage was the only parameter explaining the difference. For Evia™ DR-T and for Identity™ DR Adx, the difference between modeled survival curves could be explained by the automatic threshold management mode. Overall, the model showed a good fit for CRT-P available in the Swedish registry (InSync III™, Invive™, Frontier II™, Anthem™). This comparison could not be pursued further since the real-life programing of implants was not available on the Swedish registry website. Finally, the model showed consistency between real-life and modelled survival curves for most CIEDs from previous generations (Figure 2).

### Estimation of current devices’ longevity, using the PCI model

#### Conventional pacemakers

The aggregated survival curves showed wide differences between devices and manufacturers. Accolade™ SR and DR have the shortest estimated lifespan while the extended longevity DR version of Accolade™ offered the best longevity. The impact of programming options is reported in *Supplementary in Materials 2*.

Among possible settings, the beat-to-beat automatic threshold management algorithm (Auto capture^TM^, Automatic capture^TM^) tended to straighten product survival curve with energy savings on side of the inflexion point and energy cost on the other side, suggesting that optimal programming could extend median product longevity.

Reduction of ventricular pacing for intermittent AVB via AAI/DDD mode (available for Btk, Mcp and Mdt devices) extended median longevity for the corresponding devices and reduced the difference between Accolade™ DR EL and Azure™ XT DR or Alizea™ DR.

The number of remote transmissions had a marginal effect on longevity. Two devices (Edora™, Alizea™) showed a reduced longevity simply by activation of remote monitoring (by a one-off increase of current for Edora™ or by a reduction of battery capacity for Alizea™).

#### Leadless pacemakers

In the single chamber segment, Aveir™ V significantly outperformed Micra™ leadless pacemaker thanks to its larger battery capacity. Nevertheless, the Micra™ equipped with Capture™ management showed a survival curve at 1,5V that matched the Aveir™ survival curve at 2,5V (not equipped with Autocapture^TM^).

For two chamber leadless devices, two survival curves were considered, one for each capsule. The Aveir™ Atrial capsule had a shorter lifespan compared with the ventricular capsule. Simulation emphasizes the need to optimize pacing outputs for the atrial capsule and the importance of using the RV pacing avoidance algorithm.

#### CRT-P

Of the CRT devices, the modelled survival curves showed a significant difference between Visionist™ and similar devices. Only Percepta™ with the aCRT™ algorithm reached a similar longevity performance. Activation of MPP™ pacing negatively impacts longevity (Figure 3).

## Discussion

The current study firstly describes a novel way to estimate generator longevity by combining current and battery capacity, and then validates this model across a variety of programming options and previous generation of devices by comparing the modeled data with observed longevity from a country-wide registry, and finally provides estimations of the longevities of currently implanted devices for which there are no reliable observed data.

Longevities from user manuals are difficult to use for implant decision making because manufacturers provide these with a variety of settings, pacing options and configurations. In addition, the lack of a common framework does not facilitate an understanding of the determinants of longevity and a comparison between devices. Calculations of longevity not only should use settings reflecting clinical practice, but also split power consumption according to unavoidable current usage (background current) and optional algorithms to help practitioners in their implant decision and subsequent programming. We summarize here the key findings and a few recommendations.

### Battery capacity and background current

Battery capacity as a standalone criterion is irrelevant. This study illustrates this using the examples of leadless pacemakers which, despite much lower battery capacity, achieve a reasonable longevity compared with a standard pacemaker. On the other hand, background current plays a key role (50% of PCI is due to I_background_ across all devices). Leadless pacemakers for example benefit from a substantially lower background current compared with standard pacemakers. Therefore, both battery capacity and background current need to be combined in order to provide the foundation for longevity assessment. This is the purpose of the PCI proposed here. The index combines background current (i.e C/(365*24*10^^-6^ I_b_) and battery capacity and thereby the energy capacity (in years) available for pacing and programming options. Applying this criterion could contribute to personalized device selection, with a focus on device longevity for a wide range of patients (see table 4)

### Pacing current

The present analysis did not reveal major differences between manufacturers within each category of device, but, by exploring the differences between conventional pacemakers and leadless pacemakers, demonstrated the relative importance of energy consumption and its impact on device longevity as long as lower pacing outputs (<1.5V) can be achieved without loss of capture. As pacing output reaches 2.5V, power consumption increases and longevity is significantly impacted particularly in those devices with lower battery capacity, emphasizing the need to target pacing output close to 1-1.5V. Particularly in leadless dual chamber devices, the atrial capsule which has a modest energy capacity has the potential to limit longevity if atrial pacing is above 80% due to the additional costs of inter-capsule communication. Presently therefore, a dual chamber approach to SND requires careful consideration, especially given the younger age of the affected population. On the other hand, AV Block, with reliable intrinsic atrial activity is likely to be a more useful application for these until further technological developments improve the energetic demand of this connection.

### Current drain from optional programming

Conventional pacemakers are often chosen specifically for their programming options but the impact of these on longevity is highly variable and depends upon the manufacturer and the device generation. Amongst RVPa algorithms, AAI/DDD mode may be more effective in reducing RV pacing than other algorithms provided it can deal with intermittent AVB.

Automatic threshold management has the potential for an adverse effect on battery longevity if the threshold is low and should be deactivated in this situation. The benefit of automatic threshold management is also limited in patients where a RVPa is effective (18). ‘Daily’ algorithms are also preferable whereas beat-to-beat algorithms have a larger energy cost. For leadless pacemakers, as longevity is very sensitive to pacing outputs, daily automatic threshold management may be more critical for longevity as for standard pacemaker.

Whether extended IEGM storage and remote monitoring are required should be carefully reviewed on a patient-by-patient basis. For some devices activation of remote monitoring generates a “one off” energy cost; fortunately, the impact of the number of transmissions on longevity remains marginal.

### Overall PCI and longevity

Overall there has been an increase in longevity of more than one year for conventional SR/DR pacemakers between previous and current generation devices, although there has been a reduction of longevity for CRT-P devices. For leadless devices longevity is similar to that of a standard single chamber pacemaker, but as described, the longevity of two chamber leadless devices is currently predicted to be substantially lower than a standard dual chamber pacemaker. Disparities among suppliers still prevail and should be taken into consideration by clinicians for decision making.

### Product survival curves

Our concept was successfully validated using real-life data from previous generation devices supporting our proposal that the calculated forecasted product survival curves for current devices will be robust.

### Limitations

#### Current drain modeling

For some devices, pacing current data could not be obtained for all output values and were hypothesized by assuming similarities among same generation devices. These assumptions were mitigated via an analysis of the relationship between current and outputs highlighting consistency across devices. Options which did not specifically lead to an impact on longevity were assumed to require minimal current (e.g. RVPa algorithms and standard IEGM storage) and are not considered as generating additional energy cost.

#### Harmonization of nominal conditions and calculation of nominal PCI: validation of the model

The literature review performed to retrieve standard pacemaker parameters provided information on the average settings of devices. As RVPa were not consistently investigated via clinical trials, percentages of pacing achieved were assumed to be identical according to type (AAI/DDD mode, hysteresis mode such as SAV+^®^, VIP^®^) for SND patients. The impact of AAI/DDD modes (MVP^®^, SafeR^®^, Vp suppression^®^) for intermittent AVB available was investigated in clinical trials and considered in the model. We have also assumed that settings are stable beyond 2-3 years as we lack clinical data over a larger timeframe. The need to assume stability and permanence of success of algorithms such RVPa and aCRT is also a limitation (AV delays lengthen with time).

#### Modeling survival curves

The main limitation is the fact that real-life programming of implants was not accessible through the Swedish registry website and hence the model could not be tested with all the combination of options available. The consistency observed between real-life and modelled survival curves suggest that real-life programming does not significantly depart from the settings populated in the model, but this could not be verified.

### Conclusions

Projected longevities of CIEDs are needed for device selection and optimal programming at the time of the implant. Information from user manuals remains difficult to apply, due to a lack of harmonization in the estimated longevities provided by manufacturers regarding settings and programming conditions. We present for the first time, a model based on the Power Consumption Index (PCI) which offers the potential to compare longevity between devices under multiple settings and programming conditions. Longevities estimated by the PCI model appear to be consistent with real-life data for multiple CIED models from different manufacturers. This information could provide implanters and their patients the opportunity for personalized pacemaker hardware prescription whilst also paving the way towards standardized reporting of CIED longevity.

## Data Availability

All data produced in the present study are available upon reasonable request to the authors

## Acknowledgements

The authors thank Ernest W. Lau, MD for insightful discussion related to cardiac device implant longevity and Maxime Corneloup for programming Monte-Carlo simulations with Python™ software.

## Funding

The authors did not receive financial support from any organization for the submitted work.

## Conflict of interest

P.D. receives grants and honoraria from Medtronic, Boston Scientific, Abbott, and Microport

S.B. is a consultant for Medtronic, Boston Scientific, Microport.

JRB is an employee of Microport CRM who supervised Maxime Corneloup as an intern. KKW has received research funding from the British Heart Foundation, the National Institute for Health Research, the Medical Research Council. He has also received grants and honoraria for teaching and consultancy work from Medtronic, Cardiac Dimensions, Novartis, Abbott, BMS, Pfizer, Bayer and has received an unconditional research grant from Medtronic to support a PhD program at the University of Leeds.

## Abbreviations

CIED: Cardiac Implantable Electronic Devices
C: battery capacity
CRT-P: Cardiac resynchronization therapy with pacemaker
I: current drain
I_background_: background current
I_pacing_: pacing current
I_remote/IEGM/Algo_: current for optional settings
I_remote_: current for remote monitoring
I_IEGM_: current for IEGM storage
PCI: Power Consumption Index
RVPa: right ventricular pacing avoidance
Device manufacturers: Abt (Abbott), Btk (Biotronik), Bsc (Boston Scientific), Mdt (Medtronic), Mcp (Microport)

## Notes

### Funding Statement

This study did not receive any funding

### Summary of Updates

Some of the grammar has been improved and there is information on the source of the real world data from the Swedish Pacemaker and ICD registry.

## References

1. Schaer BA, Koller MT, Sticherling C, Altmann D, Joerg L, Osswald S. Longevity of implantable cardioverter-defibrillators, influencing factors, and comparison to industry-projected longevity. Heart Rhythm 2009;6:1737–43. doi: 10.1016/j.hrthm.2009.09.013

2. Alam MB, Munir MB, Rattan R, Adelstein E, Jain S, Saba S. Battery longevity from cardiac resynchronization therapy defibrillators: differences between manufacturers and discrepancies with published product performance reports. Europace 2017;19:421–424. doi: 10.1093/europace/euw044

3. Munawar DA, Mahajan R, Linz D, Wong GR, Khokhar KB, Thiyagarajah A, Kadhim K, Emami M, Mishima R, Elliott AD, Middeldorp ME, Roberts-Thompson KC, Young GD, Sanders P, Lau DH. Predicted longevity of contemporary cardiac implantable electronic devices: A call for industry-wide “standardized” reporting. Heart Rhythm 2018;15:1756–1763. doi: 10.1016/j.hrthm.2018.07.029

4. Boriani G, Merino J, Wright DJ, Gadler F, Schaer B, Landolina M. Battery longevity of implantable cardioverter-defibrillators and cardiac resynchronization therapy defibrillators: technical, clinical and economic aspects. An expert review paper from EHRA. Europace 2018;20:1882–1897. doi: 10.1093/europace/euy066

5. Censi F, Calcagnini G, Mattei E, Ricci RP, Zoni Berisso M, Landolina M, Boriani G. Estimate and reporting of longevity for cardiac implantable electronic devices: a proposal for standardized criteria. Expert Rev Med Devices 2021;18:1203–1208. doi: 10.1080/17434440.2021.2013199

6. Dean J, Pacemaker battery scandal. BMJ 2016;352:i228

7. Ellis CR, Dickerman DI, Orton JM, Hassan S, Good ED, Okabe T, Andriulli JA, Quan KJ, Greenspon AJ. Ampere Hour as a Predictor of Cardiac Resynchronization Defibrillator Pulse Generator Battery Longevity: A Multicenter Study. Pacing Clin Electrophysiol 2016;39:658–68. doi: 10.1111/pace.12831

8. Lau EW. Technologies for prolonging cardiac implantable electronic device longevity. Pacing Clin Electrophysiol 2017;40:75–96

9. Paton MF, Landolina M, Billuart JR, Field D, Sibley J, Witte K. Projected longevities of cardiac implantable defibrillators: a retrospective analysis over the period 2007-17 and the impact of technological factors in determining longevity. Europace 2020;22:149–155. doi: 10.1093/europace/euz222. Erratum in: Europace. 2020 Jan 1;22(1):155. doi: 10.1093/europace/euz308

10. Evans JM, Cleves A, Morgan H, Millar L, Carolan-Rees G. ENDURALIFE-Powered Cardiac Resynchronisation Therapy Defibrillator Devices for Treating Heart Failure: A NICE Medical Technology Guidance. Appl Health Econ Health Policy 2018;16:177–186. doi: 10.1007/s40258-017-0354-6

11. Lau EW. Longevity decoded: Insights from power consumption analyses into device construction and their clinical implications. Pacing Clin Electrophysiol 2019;42:407–422. doi: 10.1111/pace.13642

12. Boston Scientific Longevity calculator, https://www.bostonscientific.com/en-EU/medical-specialties/electrophysiology/device-longevity/longevity-calculator.disclaimer.html

13. Swedish ICD, Pacemaker Registry, https://www.pacemakerregistret.se/icdpmr/start.do

14. Udo EO, van Hemel NM, Zuithoff NP, Barrett MJ, Ruiter JH, Doevendans PA, Moons KG. Incidence and predictors of pacemaker reprogramming: potential consequences for remote follow-up. Europace 2013;15:978–83. doi: 10.1093/europace/eut002

15. Auricchio A, Ellenbogen KA. Reducing Ventricular Pacing Frequency in Patients With Atrioventricular Block: Is It Time to Change the Current Pacing Paradigm? Circ Arrhythm Electrophysiol 2016;9:e004404. doi: 10.1161/CIRCEP.116.004404

16. Stockburger M, Defaye P, Boveda S, Stancak B, Lazarus A, Sipötz J, Nardi S, Rolando M, Moreno J. Safety and efficiency of ventricular pacing prevention with an AAI-DDD changeover mode in patients with sinus node disease or atrioventricular block: impact on battery longevity-a sub-study of the ANSWER trial. Europace 2016;18:739–46. doi: 10.1093/europace/euv358

17. Biffi M, Melissano D, Rossi P, Kaliska G, Havliĉek A, Pelargonio G, Romero R, Guastaferro C, Menichelli M, Vireca E, Frisoni J, Boriani G, Malacky T. The OPTI-MIND study: a prospective, observational study of pacemaker patients according to pacing modality and primary indications. Europace 2014;16:689–97. doi: 10.1093/europace/eut387

18. Benezet-Mazuecos J, Iglesias JA, Rubio JM, Cortés M, de la Cruz E, de la Vieja JJ, Calle S, Farré J. Limitations of the AutoCapture™ Pacing System in patients with cardiac stimulation devices. Europace 2014;16:1469–75. doi: 10.1093/europace/euu080

19. Pierantozzi A, Landolina M, Agricola T, Lunati M, Pisanò E, Lonardi G, Bardelli G, Proclemer A, Speca G, Zucchi G, Marseglia A, Valsecchi S, Bocconcelli P; Italian Clinical Service Left Ventricular Capture Management Group. Automatic adjustment of stimulation output in resynchronization therapy: impact and effectiveness in clinical practice. Europace 2011;13:1311–8. doi: 10.1093/europace/eur118

20. Zoppo F, Gagno G. Left ventricle automatic pacing threshold management in CRT systems: A comprehensive review. J Cardiovasc Electrophysiol 2020;31:2489–2498. doi: 10.1111/jce.14630

21. Martin DO, Lemke B, Birnie D, Krum H, Lee KL, Aonuma K, Gasparini M, Starling RC, Milasinovic G, Rogers T, Sambelashvili A, Gorcsan J 3rd, Houmsse M; Adaptive CRT Study Investigators. Investigation of a novel algorithm for synchronized left-ventricular pacing and ambulatory optimization of cardiac resynchronization therapy: results of the adaptive CRT trial. Heart Rhythm 2012;9:1807–14. doi: 10.1016/j.hrthm.2012.07.009

22. Reynolds D, Duray GZ, Omar R, Soejima K, Neuzil P, Zhang S, Narasimhan C, Steinwender C, Brugada J, Lloyd M, Roberts PR, Sagi V, Hummel J, Bongiorni MG, Knops RE, Ellis CR, Gornick CC, Bernabei MA, Laager V, Stromberg K, Williams ER, Hudnall JH, Ritter P; Micra Transcatheter Pacing Study Group. A Leadless Intracardiac Transcatheter Pacing System. N Engl J Med 2016;374:533–41. doi: 10.1056/NEJMoa1511643

23. Piccini JP, Stromberg K, Jackson KP, Laager V, Duray GZ, El-Chami M, Ellis CR, Hummel J, Jones DR, Kowal RC, Narasimhan C, Omar R, Ritter P, Roberts PR, Soejima K, Zhang S, Reynolds D; Micra Transcatheter Pacing Study Group. Long-term outcomes in leadless Micra transcatheter pacemakers with elevated thresholds at implantation: Results from the Micra Transcatheter Pacing System Global Clinical Trial. Heart Rhythm 2017;14:685–691. doi: 10.1016/j.hrthm.2017.01.026

24. Knops RE, Reddy VY, Ip JE, Doshi R, Exner DV, Defaye P, Canby R, Bongiorni MG, Shoda M, Hindricks G, Neužil P, Rashtian M, Breeman KTN, Nevo JR, Ganz L, Hubbard C, Cantillon DJ; Aveir DR i2i Study Investigators. A Dual-Chamber Leadless Pacemaker. N Engl J Med 2023;388:2360–2370. doi: 10.1056/NEJMoa2300080

25. Shantha G, Brock J, Singleton MJ, Schmitt AJ, Kozak P, Bodziock G, Bradford N, Whalen P, Bhave P. A comparative study of the two leadless pacemakers in clinical practice. J Cardiovasc Electrophysiol 2023;34:1896–1903. doi: 10.1111/jce.16019

