## Supplemental material 2 for "Cardiac Implantable Electronic Devices’ longevity: A novel modelling tool for estimation and comparison" (from another version of this preprint: version 1, 2025-03-31)

**Supplementary Materials 2**

1. **Power consumption and nominal longevities**

Based on pacing current formulas and the current needed for each configuration, the PCI and the nominal longevity can be estimated for each device (see table below) under the following assumptions:

- For conventional pacemakers, the settings considered were: basic rate of 60bpm, pacing output at 2.5V, pulse duration: 0.4ms and impedance: 500 ohms for both A&V. For VVI pacemakers, ventricular pacing: 90% and for dual chamber pacemakers, atrial pacing: 70% for SND, 30% for AVB (51% on average) and ventricular pacing assumptions accounted for the difference between AAI/DDD mode and other RVP algorithms (29% vs 47%).

Options such as sensor, IEGM storage and remote (2 transmissions/year) are reported in Figure 2.

- For leadless VVI and DDD pacemaker, the settings used were: basic rate: 60bpm, pulse duration: 0.25ms for Micra™ and 0.4 ms for Aveir™, impedance: ~600 ohms for ventricle and ~300 ohms for the atrium. Pacing outputs were not reported in studies and two options were considered: 1.5V or 2.5V reflecting the level of confidence of practitioners in adapting output (thresholds observed were typically low: 1.25V at implant and 0.75V weeks after). Pacing percentages were the same as for conventional pacemakers. Hysteresis mode was applied for DDD.
- For CRT-P, BIV was the standard pacing mode with alternative options such as aCRT^TM^ or MPP^TM^ pacing.

**PCI and nominal longevities**

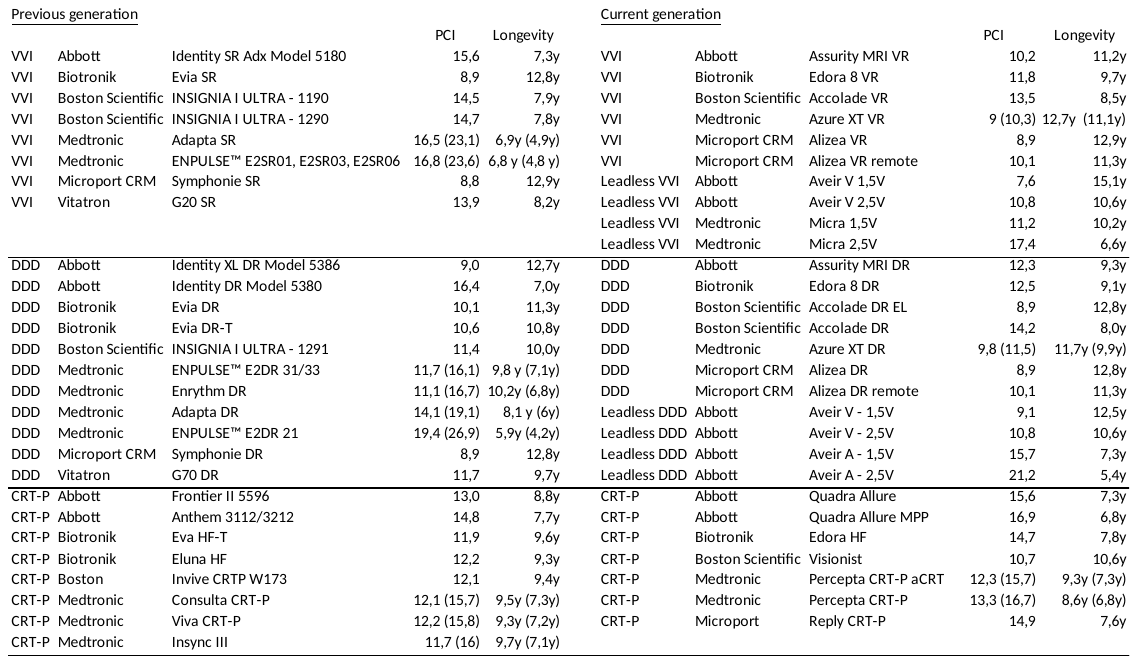

**Evolution**

|  | | |  | | | Power consumption index split (PCI) | | | Longevity (years) | | |
| --- | --- | --- | --- | --- | --- | --- | --- | --- | --- | --- | --- |
|  | Generation | Background | Pacing | Sensor | Ext. IEGM | FUP | Remote | Total wo ext. IEGM | Total  w  ext. IEGM | wo ext. IEGM | w.  ext. IEGM |
| VVI | Previous | 10,4 | 2,4 | 0,8 | 1,7 | 0,1 | 0,0 | 13,7 | 15,4 | 8,8y | 8,3y |
| VVI | New | 6,9 | 1,9 | 0,8 | 0,2 | 0,1 | 0,9 | 10,6 | 10,8 | 11,0y | 10,8y |
| DDD | Previous | 9,4 | 1,8 | 0,7 | 2,1 | 0,1 | 0,2 | 12,2 | 14,3 | 9,9y | 9,0y |
| DDD | New | 7,6 | 1,7 | 0,7 | 0,2 | 0,1 | 0,8 | 11,0 | 11,2 | 10,7y | 10,5y |
| CRT-P | Previous | 7,9 | 3,8 | 0,7 | 1,4 | 0,1 | 0,0 | 12,5 | 13,9 | 9,2y | 8,3y |
| CRT-P | New | 7,8 | 4,3 | 0,8 | 1,0 | 0,1 | 1,1 | 14,1 | 15,0 | 8,3y | 7,8y |

2. **Survival curves generated by the Monte-Carlo modelization**

**Settings and distribution used for Monte-Carlo simulations**

**Conventional single and dual pacemakers**

|  | **SND** | **Inter AVB** | **AVB perm** | **Distribution** |
| --- | --- | --- | --- | --- |
| V lead Z (Ω) | 500 ± 120 | 500 ± 120 | 500 ± 120 | Normal |
| V width (ms) | 0.35 [0.35; 0.35] | 0.35 [0.35; 0.35] | 0.35 [0.35; 0.35] | Constant |
| V output (V) | 2.5 [2.5; 3.5] | 2.5 [2.5; 3.5] | 2.5 [2.5; 3.5] | Normal |
| V output-LBBAP (V) | 2x 0,7 ± 0,24 | 2 x 0,7 ± 0,24 | 2x 0,7 ± 0,24 | Normal |
| VP VVI (%) | 90 [83; 100] | 90 [83; 100] | 90 [83; 100] | Normal |
| VP AAI/DDD* (%) | 2.2 [0.0; 29.9] | 53.5 [1.7; 95.9] | 89.2 [24.0; 99.3] | Per quartile |
| VP Other RVP* (%) | 2.2 [0.0; 29.9] | 97.9 [81.0; 99.6] | 98.2 [83.4; 99.6] | Per quartile |
| A lead Z *(Ω) | 450 ± 120 | 450 ± 120 | 450 ± 120 | Normal |
| A width *(ms) | 0.35 [0.35; 0.35] | 0.35 [0.35; 0.35] | 0.35 [0.35; 0.35] | Constant |
| A output *(V) | 2.5 [2.5; 3.5] | 2.5 [2.5; 3.5] | 2.5 [2.5; 3.5] | Normal |
| AP* (%) | 70.1 [30.5; 92.0] | 27.5 [9.0; 64.5] | 43.7 [2.6; 73.5] | Per quartile |

Legend : median [Q1; Q3], * specific to dual chamber pacemaker / pacing outputs are not reported yet for LBBAP pacing and are assumed to be twice as high as threshold value.

**Single and dual chamber leadless pacemakers**

|  | **SND** | **Inter AVB** | **AVB perm** | **Distribution** |
| --- | --- | --- | --- | --- |
| V lead Z (Ω) | 627± 100 | 627± 100 | 627± 100 | Normal |
| V width (ms) | 0,24 ^M^ / 0,4 ^A^ | 0,24 ^M^ / 0,4 ^A^ | 0,24 ^M^ / 0,4 ^A^ | Constant |
| V output (V) | 2.5 [2.5; 3.5]  or 2x 0,75±0,5* | 2.5 [2.5; 3.5]  or 2x0,75 ±0,5* | 2.5 [2.5; 3.5]  or 2x0,75± 0,5* | Normal |
| VP VVI (%) | 90 [83; 100] | 90 [83; 100] | 90 [83; 100] | Normal |
| VP RVP* (%) | 2.2 [0.0; 29.9] | 97.9 [81.0; 99.6] | 98.2 [83.4; 99.6] | Per quartile |
| A lead Z (Ω)* | 315 ± 5 ^A^ | 315 ± 5 ^A^ | 315 ± 5 ^A^ | Normal |
| A width (ms)* | 0,4 ^A^ | 0,4 ^A^ | 0,4 ^A^ | Constant |
| A output (V)* | 2.5 [2.5; 3.5]  or 2x 0,75±0,5* | 2.5 [2.5; 3.5]  or 2x 0,75±0,5* | 2.5 [2.5; 3.5]  or 2x 0,75±0,5^*^ | Normal |
| AP (%)* | 70.1 [30.5; 92.0] | 27.5 [9.0; 64.5] | 43.7 [2.6; 73.5] | Per quartile |

Legend : median [Q1; Q3], * specific to dual chamber pacemaker (RVP is programmed with AV hysteresis mode), pacing conditions are selected according to clinical review for each leadless (^M^: Micra, ^A^ Aveir). See references : 17, 18, 19, 20. Comment: pacing thresholds are usually low 0.82 ± 0.8 (atrial) and 0.6 ± 0.5 (ventricle). Clinical studies do not report pacing outputs; hence two pacing output scenarios are considered.

**Cardiac resynchronisation therapy pacemakers**

|  |  | **Distribution** |
| --- | --- | --- |
| V lead Z (Ω) | 500 ± 120 | Normal |
| V width (ms) | 0.35 [0.35; 0.35] | Constant |
| V output (V) | 2.5 [2.5; 3.5] | Normal |
| VP BIV (%) | 95.1% ± 10.5% | Normal |
| VP Adapt CRT(%)* | 50.9% ± 15% | Normal |
| A lead Z (Ω) | 450 ± 120 | Normal |
| A width (ms) | 0.35 [0.35; 0.35] | Constant |
| A output (V) | 2.5 [2.5; 3.5] | Normal |
| AP (%) | 50% ± 15% | Per quartile |
| LV lead Z (Ω) | 745±325 | Normal |
| LV width (ms) | 0.35 [0.35; 0.35] | Constant |
| LV output (V) | 2.5 [1.7; 3.3] | Normal |
| LV BIV (%) | 95.1% ± 10.5% | Per quartile |

Same conditions as for conventional PM for A et V; LV conditions are extracted from literature (and specifically from Adaptive CRT trial for (*))

**Survival curves generated for previous generation devices**

| **VVI** | **DDD(R)** |
| --- | --- |
| 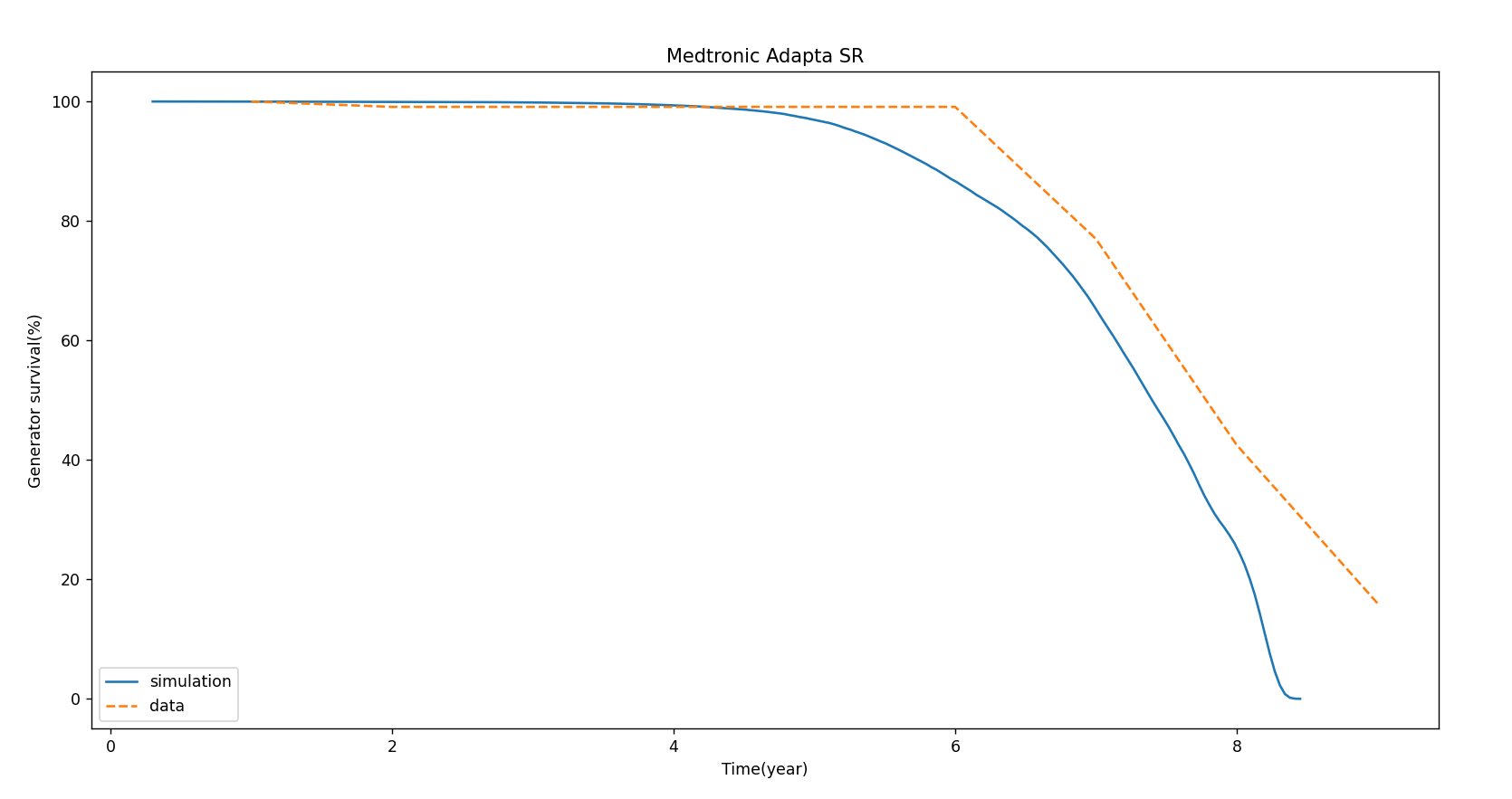 | 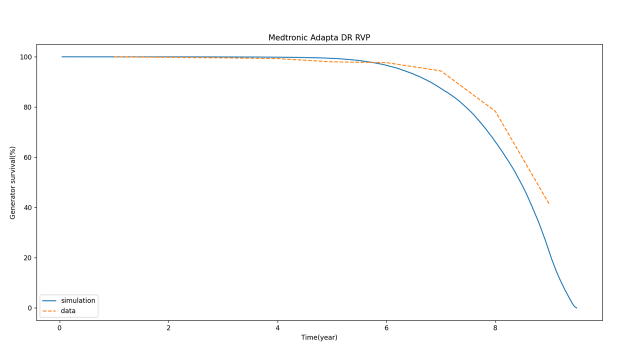 |
| 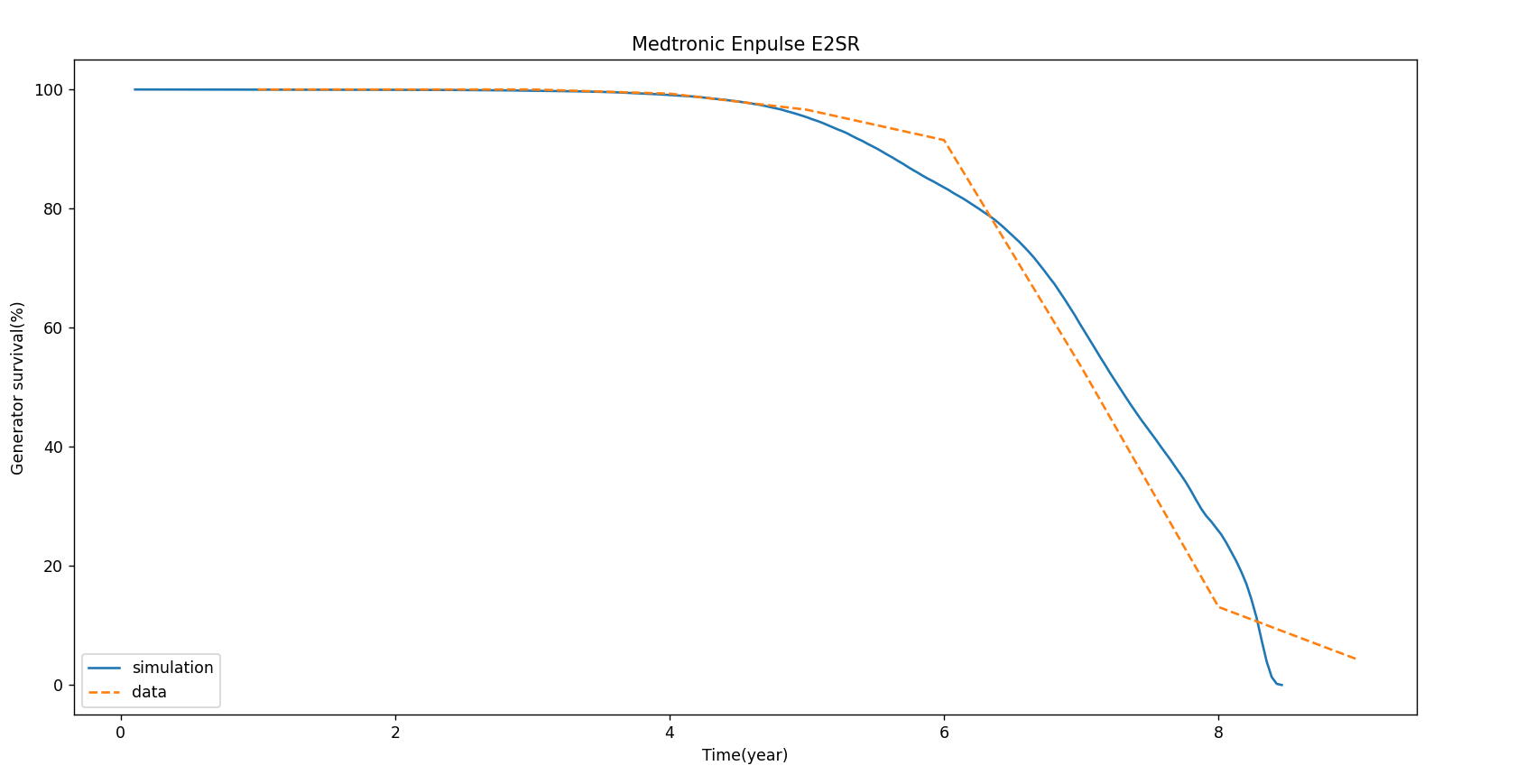 | 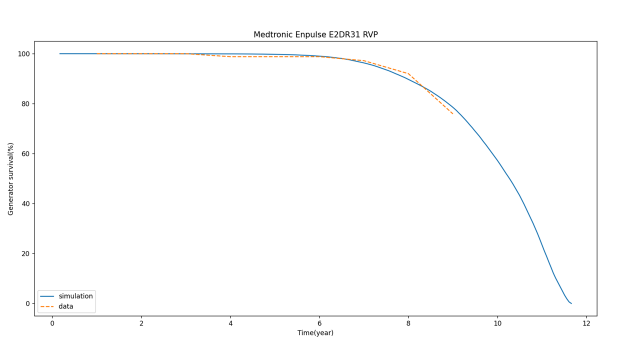 |
| 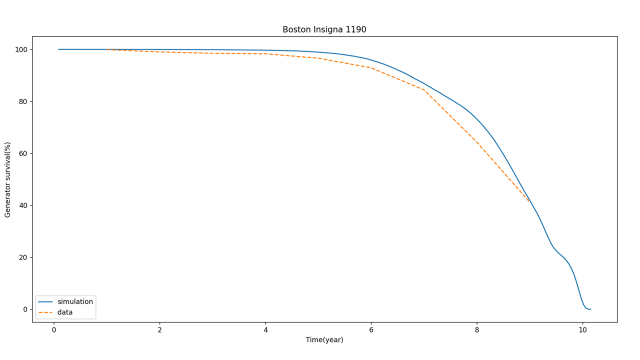 | 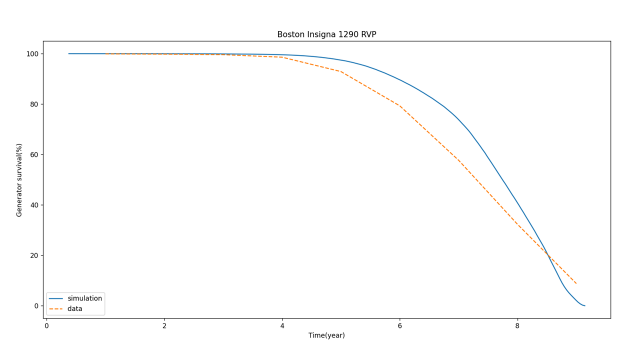 |
| 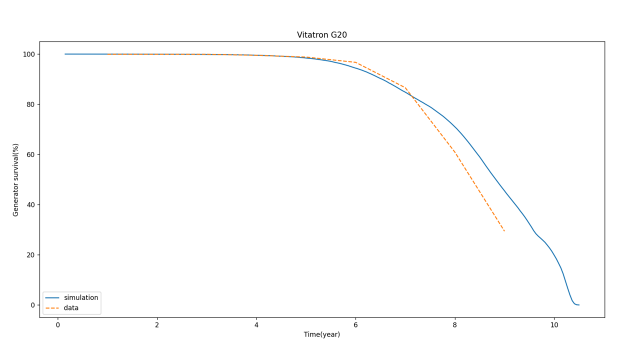 | 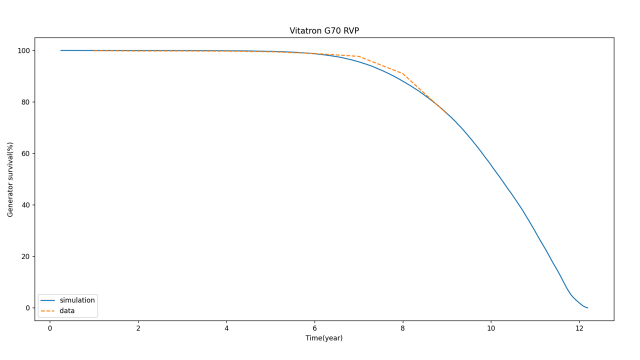 |
| 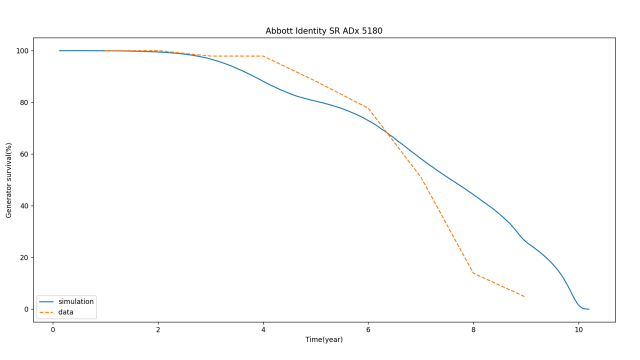 | 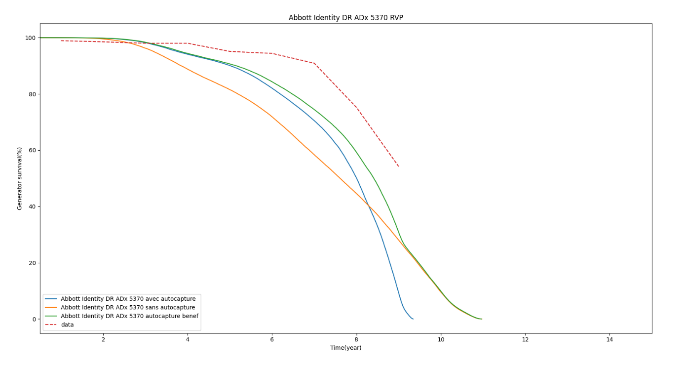 |

| **VVI** | **DDD(R)** |
| --- | --- |
| **NA** | 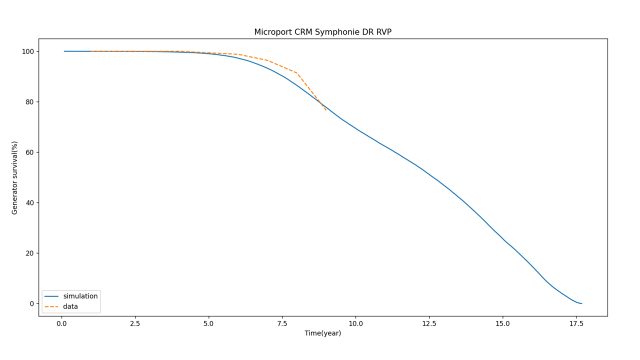 |

**Impact of settings for previous generation devices**

| **Comments** | **DDD(R)** |
| --- | --- |
| With standard settings, the modeled survival curve for the Medtronic Enrythm DR with RV pacing avoidance activated significantly deviates from all real life data. The only source of explanation is the use of IEGM storage (simulation assumes 1/3 pts without IEGM, 1/3 with IEGM, 1/3 with extended IEGM) | 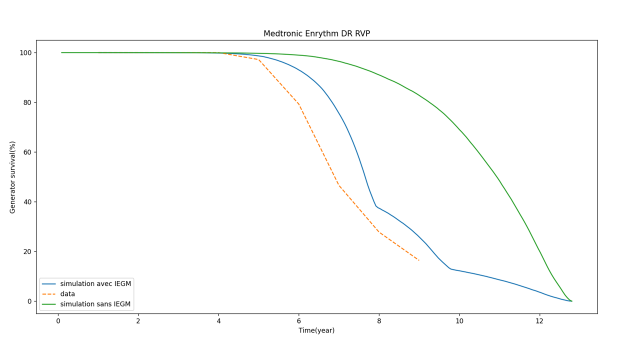 |
| With standard settings, the modeled survival curve of the Biotronik Evia DR-T deviates from real life data. The modelled survival curve with automatic threshold management on, matches real life data well. | 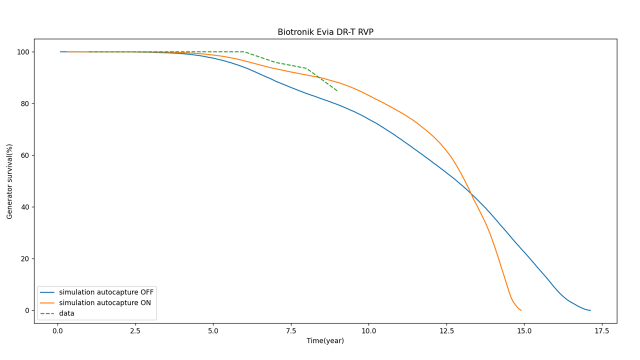 |
| With standard settings, the modeled survival curve deviates from real life data and with Auto capture^TM^ activated better matches the Swedish registry data.  The chart reports 2 scenarios (Auto capture™ activated for all patients, Auto capture™ activated only if it improves longevity – i.e. the reduction of current drain via lower output was superior to the energy cost of Auto capture™) | 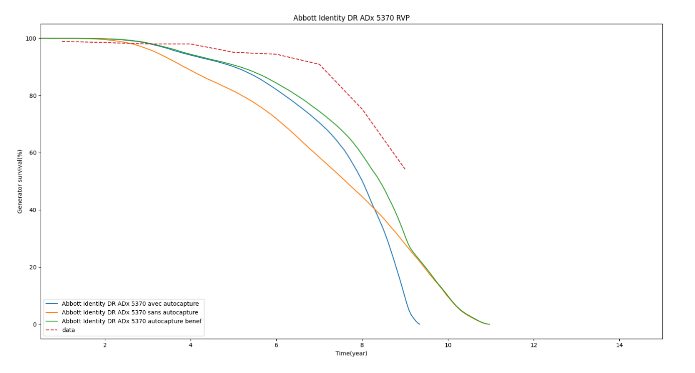 |

**Survival curves generated for CRT-P devices**

| **LV 2.5 +/-0.8V** 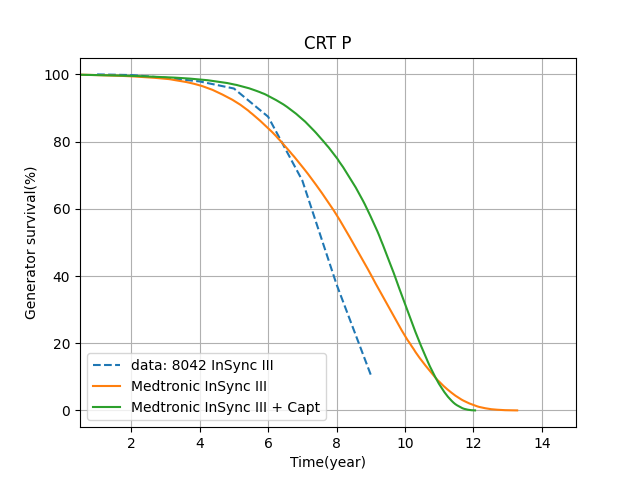 | **LV 3.5 +/-0.8V** 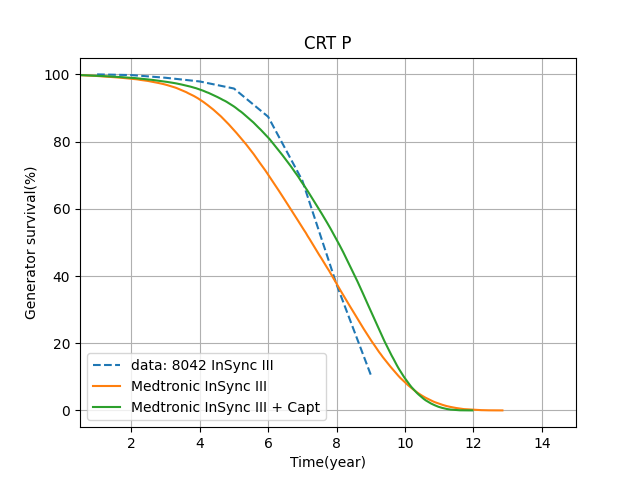 |
| --- | --- |

| 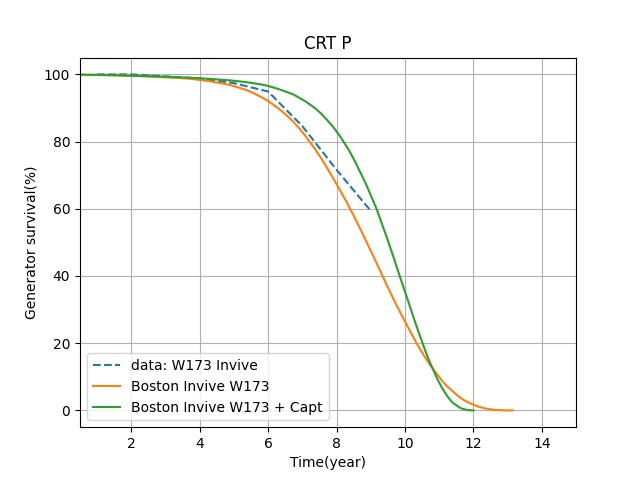 | 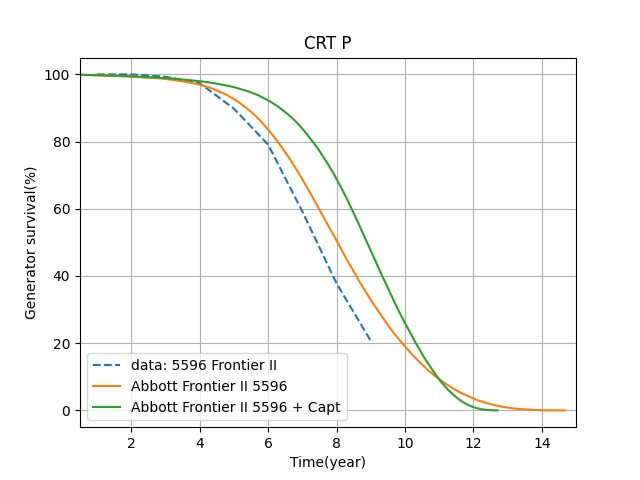 |
| --- | --- |
| 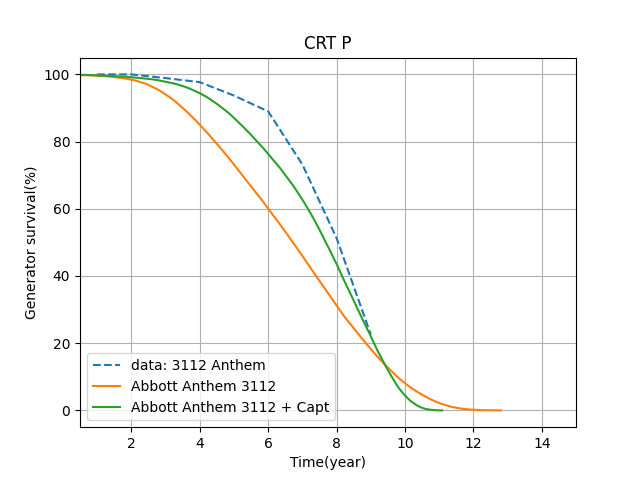 |  |

**Impact of settings for current generation devices**

**Impact of Automatic capture^TM^ on longevity**

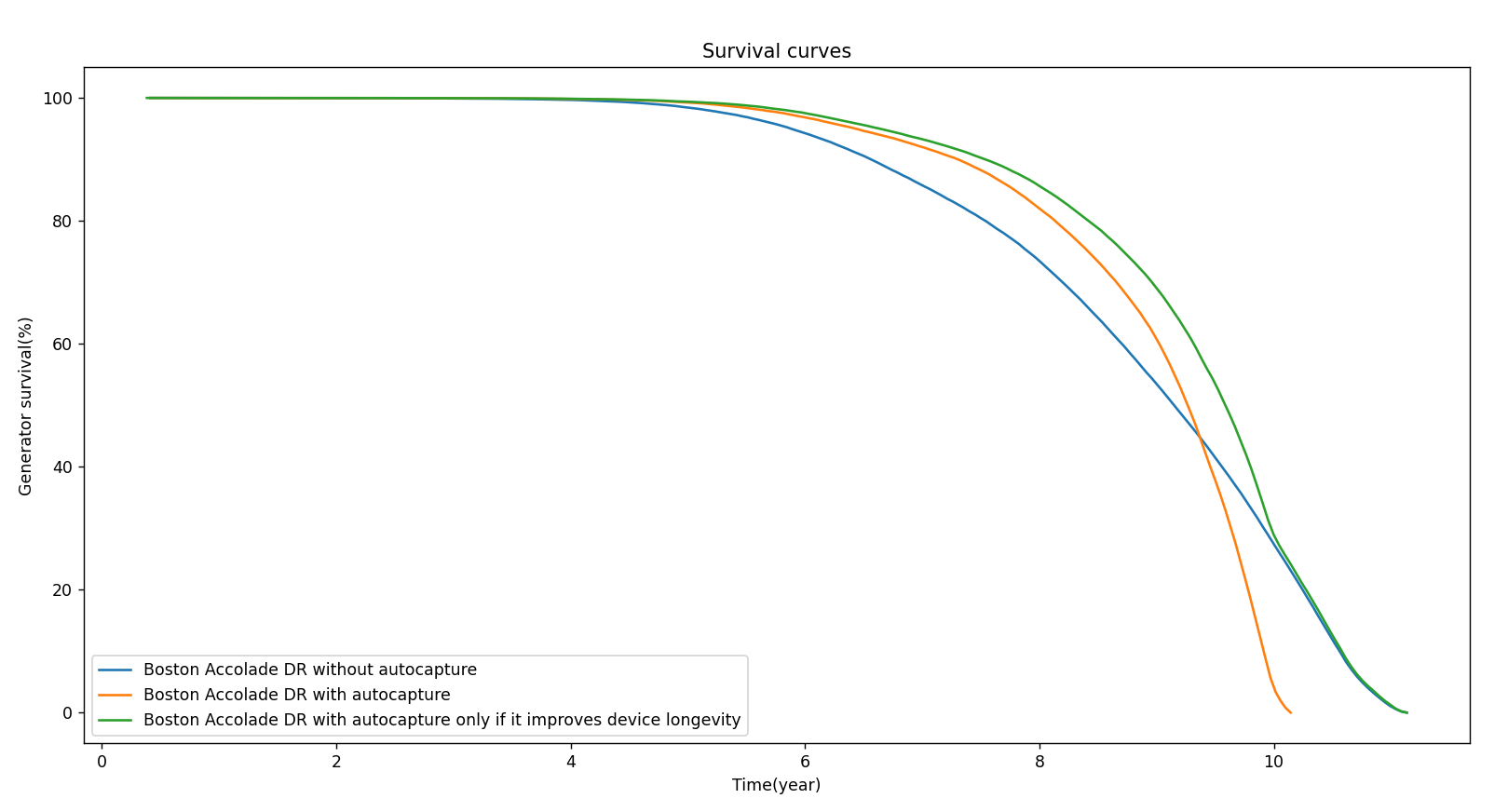

Assuming 1 μA energy cost for the algorithm and an ideal output of 1 V.

**Impact of RVP algorithm on longevity:**

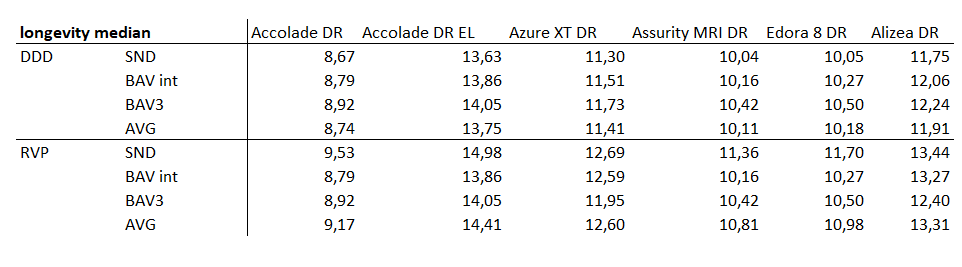

**Median longevity is derived from the simulation of survival curve according to the previously described method.**

**For intermittent AVB and AVB 3, RVP is only applied for those populations for which there is evidence of reduction of VP (MVP^TM^ ,Safe^TM^) and by extension to similar AAI/DDD mode (Btk).**

**Impact of remote monitoring and downloads**

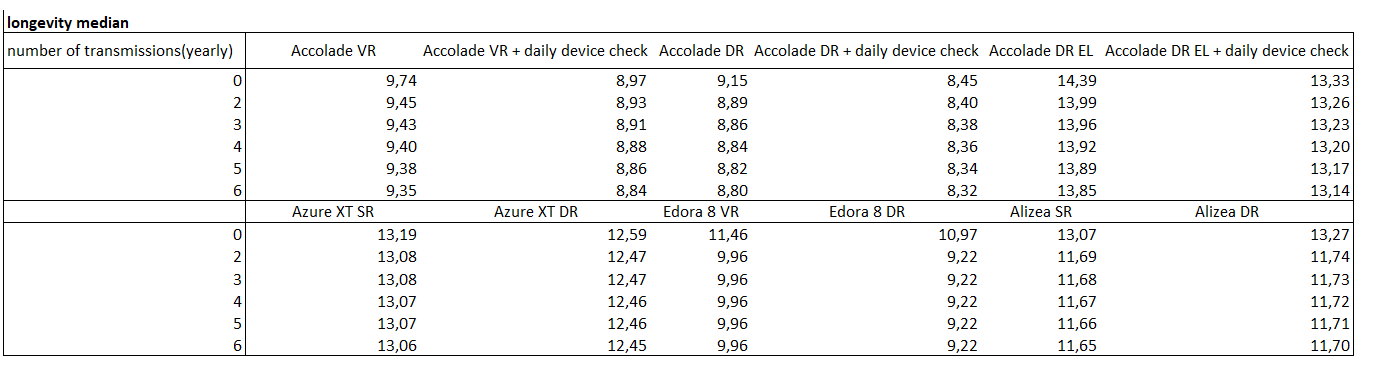

**Standard settings (see settings used for MC simulation)+activation of remote**
