## Supplemental material 1 for "Cardiac Implantable Electronic Devices’ longevity: A novel modelling tool for estimation and comparison" (from another version of this preprint: version 1, 2025-03-31)

**Supplementary Materials 1**

**Model development**

**1. Pacing current formula and regression**

**Longevity and current**

The first step was to calculate estimated energy consumption based on information from user manuals using the longevity formula L=C / (365*24*10^^-6^ I) (where h L: longevity of the device in years, C: battery capacity at ERI in Ah and I: current drain in μA) which we converted to I=C/L.

The nominal current I is the sum (I = I_background_ + I_pacing_ + I_remote_ + I_IEGM_ + I_sensor_) of all currents used to maintain the device (I_background_), provide pacing (I_pacing_) and support optional features (remote monitoring and downloads, intracardiac electrocardiogram storage and rate-response sensor).

In all user manuals, longevity is provided with different combination of pacing parameters influencing I_pacing_, while other options (remote monitoring, IEGM, sensors) are either turned on or off. These values allow calculating I for different settings.

The current I_background_ and the I_pacing_ parameters were derived by regression from the set of current values at different settings (impedance, pulse, pacing outputs). The formula used for regression is “universal” and is based on the “Pacing current formula” (Mark W. Sweesy, Fundamental electrical relationships, Feb 2009, Cardio rhythm 2009).

**Pacing current formula**

The current drain for pacing, at the electrode (I_stim_) is the same for all devices. This current drain is not delivered directly by the battery but by an electronic circuit (specific to each manufacturer) and conveyed by a cardiac lead.

The electronic circuit drain is however based upon current I_pacing_ from the battery. The efficiency of the electronic circuit is characterized by a pacing energy coefficient R_vs_ (%) which reflects the amount of energy from the battery needed to provide the required energy at the electrode. A high R_vs_ corresponds to a lower current drain from the battery and hence more efficiency.

The power needed at the electrode (V_stim_ x I_stim_) results from the multiplication of this converting factor, the pacing energy coefficient R_vs_ multiplied by the power drained at the level of the battery (V_b_ x I_pacing_).

From this first step, I_pacing_ from the battery was derived (see chart). From I_stim_ to I_pacing_ a converting factor was applied. It depends on the ratio V_stim_/V_battery_ and the ratio (1/R_vs_).

| 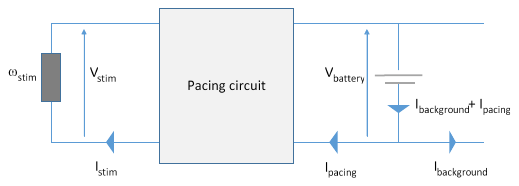 | 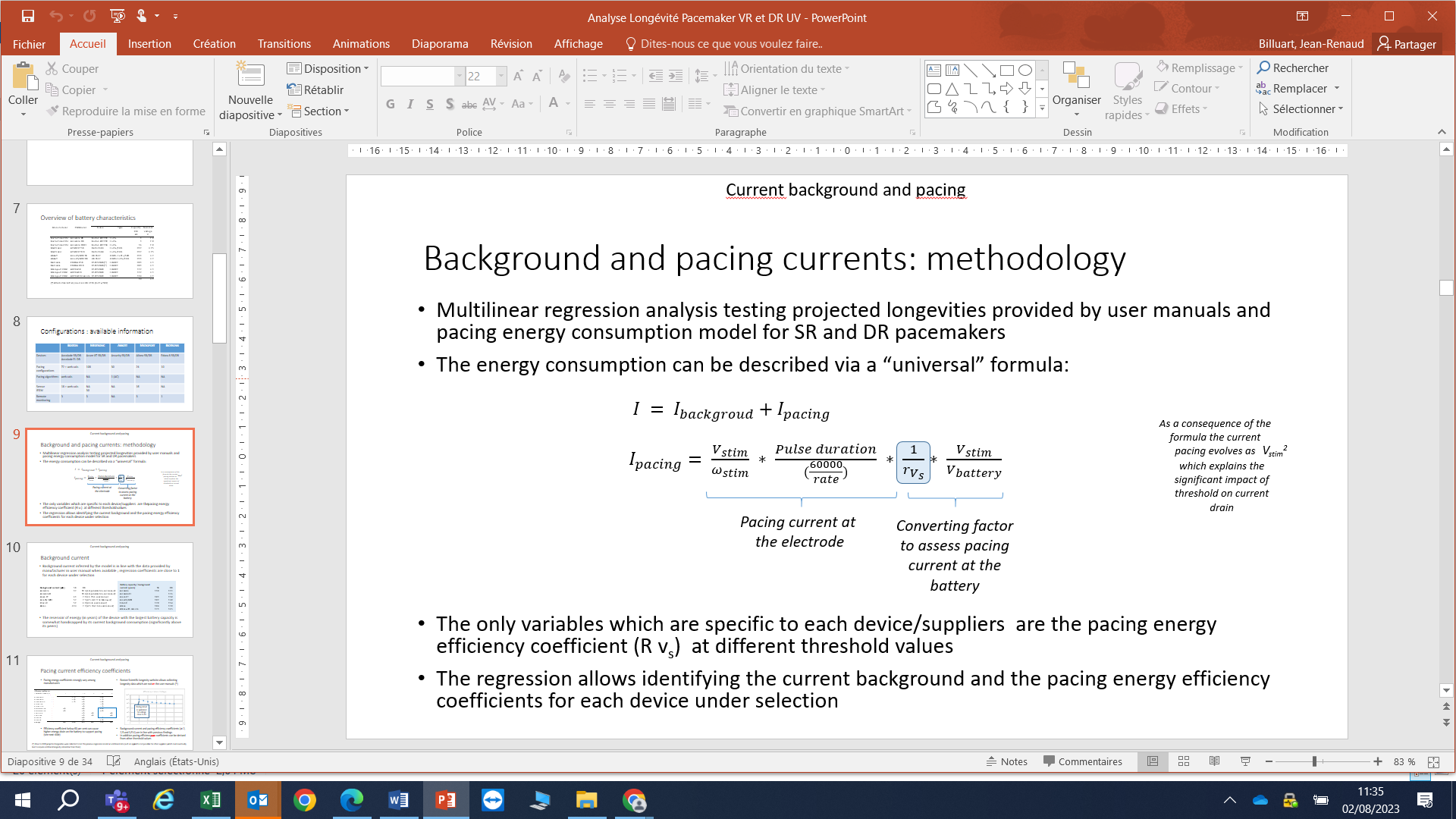 |
| --- | --- |

Pacing current depends on the square of V_stim_ which explains why pacing output is so important in energy consumption.

We assumed that the voltage at the battery level V_battery_ is constant. This is a proxy, as battery discharge changes the voltage of the battery. The behavior of the battery is specific to each supplier and not disclosed. We investigated via the Bsc longevity calculator whether this assumption could be valid for longevity calculation for Accolade SR™ and Accolade DR EL™ (see chart) and battery longevity voltage was close to the 2.8 V declared nominal longevity.

| 1000 configurations | 45000 configurations |
| --- | --- |
| 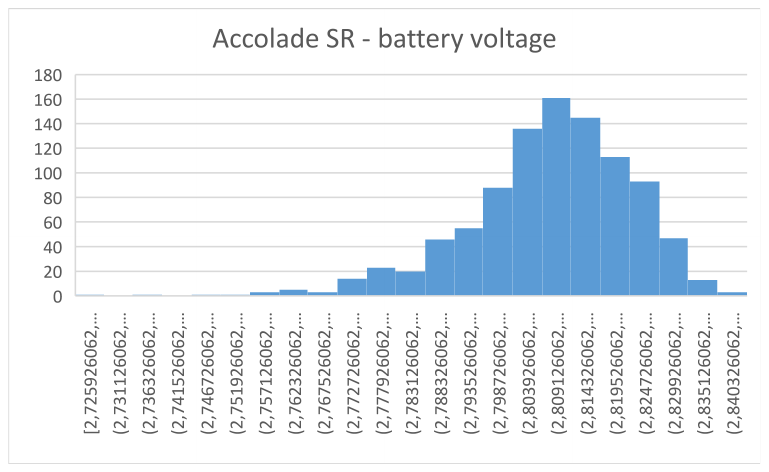 | 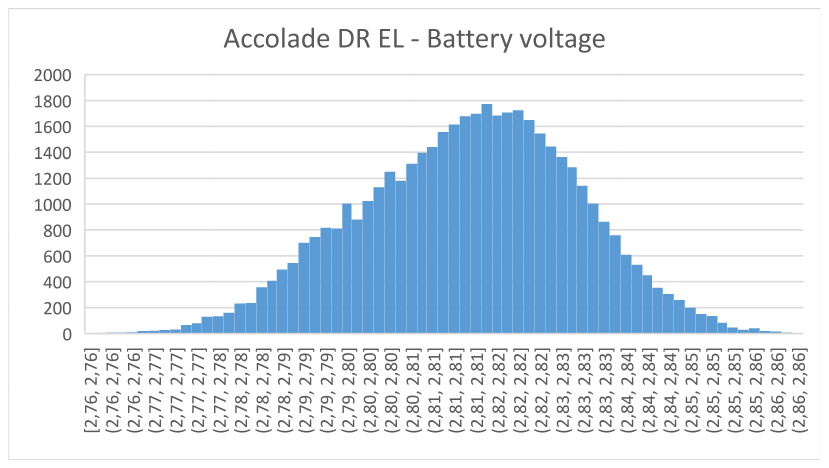 |

Note : Bsc longevity calculator provides the power consumption (Watt) and longevity (years). From longevity, the total current drain I can be derived via the formula I = C /L. The power consumption divided by the total current drain gives the battery voltage used for longevity calculation.

In the analysis, if user manuals provided sufficient data, I_background_ and R_vs_ were derived from regression and compared with user manual information. Otherwise, the value of I_background_ was taken from user manuals; pacing energy coefficients were derived from regression.

The Bsc longevity calculator allow an estimation of longevity on a broader range of parameters. Results derived from the user manual were compared with those derived from the longevity calculator. For other manufacturers, the information provided in user manuals was sometimes heterogeneous, potentially impacting the robustness of the energy model. This constitutes an intrinsic limitation of the proposed approach.

**Regression methodology**

The example below illustrates the method used for VVI pacemaker.

| **Regression analysis (Accolade SR example)** | |
| --- | --- |
| Based on the user manual or longevity website, a linear regression test was performed between two variables: the total current I (derived via the formula I = C/L) and the pacing current (derived via the pacing current formula). Indeed, longevity is provided by suppliers’ source of data and corresponding conditions. The regression allows the assessment of I_background_ and pacing energy coefficient (R_vs_) (the slope in the chart is 1/ R_vs_). | 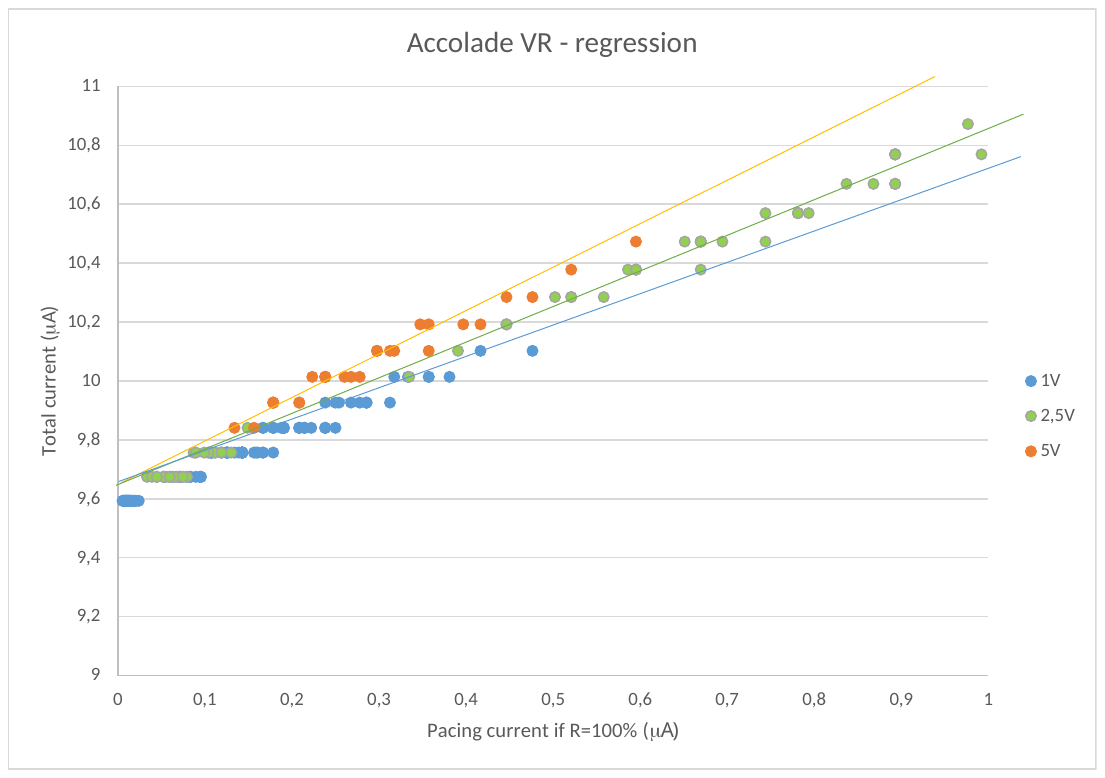 |

For dual chamber and CRT-P devices, a multilinear model was used to model pacing current for each chamber.

Where user manual provided insufficient data for a specific model, regression was conducted while testing SR and DR simultaneously or groups of devices known to share the same platform and battery technology.

**2. Results of regression and pacing efficiency coefficients**

The background currents derived from regression were reported in the core of the article and the pacing efficiency coefficients are reported in this Appendix.

For **conventional pacemakers**, regression analysis allowed the modeling of longevity with reasonable accuracy for each product. The difference between reported longevity (as per manufacturers’ manual) and modeled longevity across all manufacturers and devices reached 0.1 ±4% for previous generation devices and -0.1 ±0.7% for new generation devices (see error distribution below). For devices with a variety of configurations (Medtronic and Boston Scientific via their longevity calculator website), regression coefficient (R^2^) exceeded 90%, for all configurations.

Pacing efficiency derived from the model on average did not change between previous and current generation devices (83% vs 81% at 2.5V and 74% vs 77% at 3.5V). In previous generation devices, only one supplier (Medtronic) provided a consistent pacing coefficient above 78-80% for both 2.5V and 3.5V. In new generation devices, this objective was achieved by two suppliers (Medtronic and Boston Scentific). The analysis based on data extracted from the Boston Scientific longevity calculator website validated both the pacing coefficient derived for Accolade™, at 2.5V and 3.5V and the pacing coefficient for other threshold values. Pacing coefficients for CRT-P were on average higher (90% at 2.5V and 81% at 3.5V for implants of the new generation) which may reflect a strategy to optimize pacing current drain for CRT therapy which is more demanding on battery longevity than a standard pacemaker.

For **leadless pacemakers**, outputs reported were mainly at 1.25V and 2.5V and pacing coefficients were on average 59% and 77%)

**Conventional pacemakers**

| **Modeled longevity vs. nominal longevity provided by manuals** | |
| --- | --- |
| Previous generation 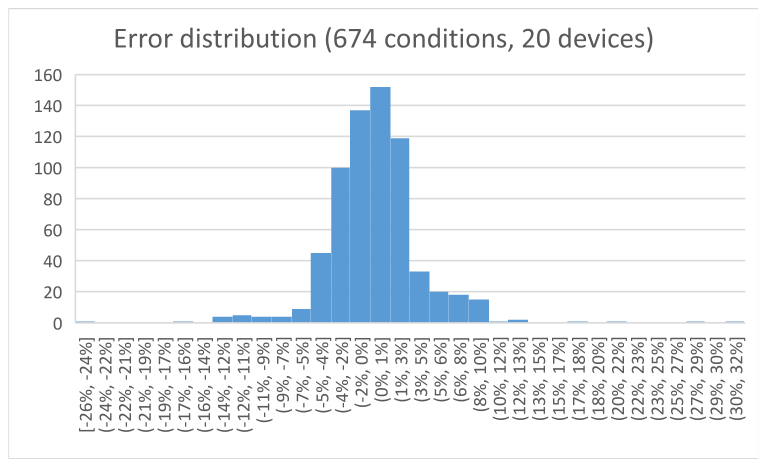 | Current generation 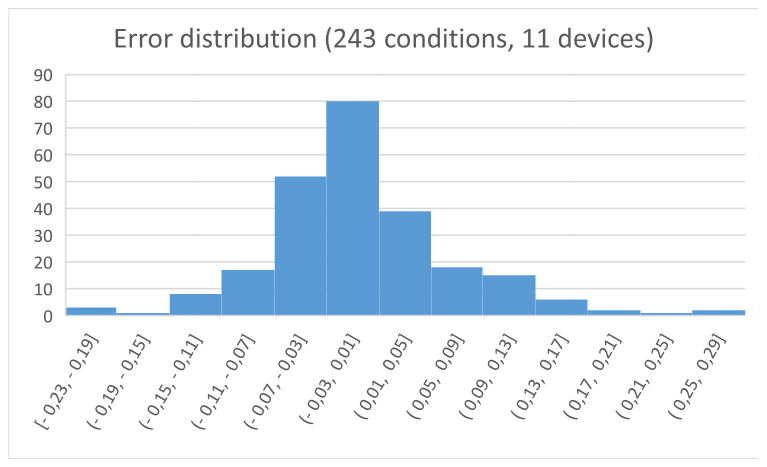 |

| 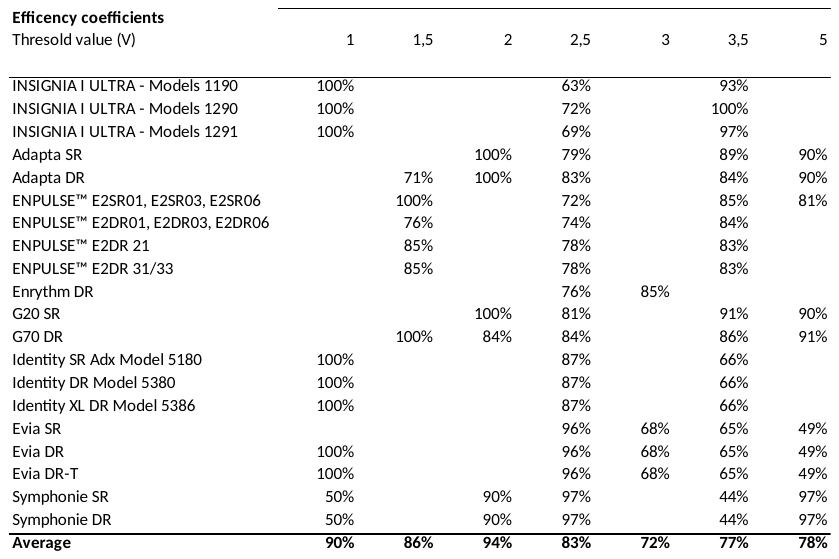 | 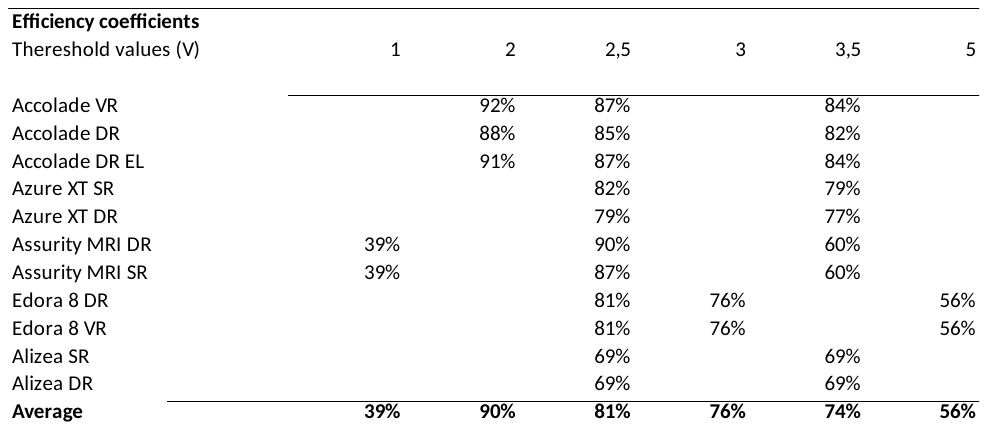 |
| --- | --- |

| **Accolade™ SR/DR pacing coefficients** 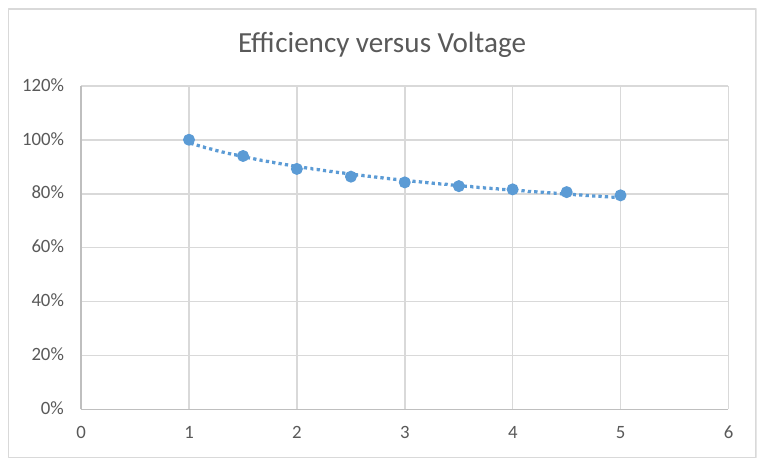 | Close to 1000 projected longevities were collected from the Boston Scientific longevity website to test regression model (R^2^: 95%) on additional data and voltages which were not provided by the user manual. Background current and pacing coefficient were subsequently derived from regression. |
| --- | --- |

**Leadless pacemakers**

Modeled longevity *vs*. nominal longevity provided by manuals

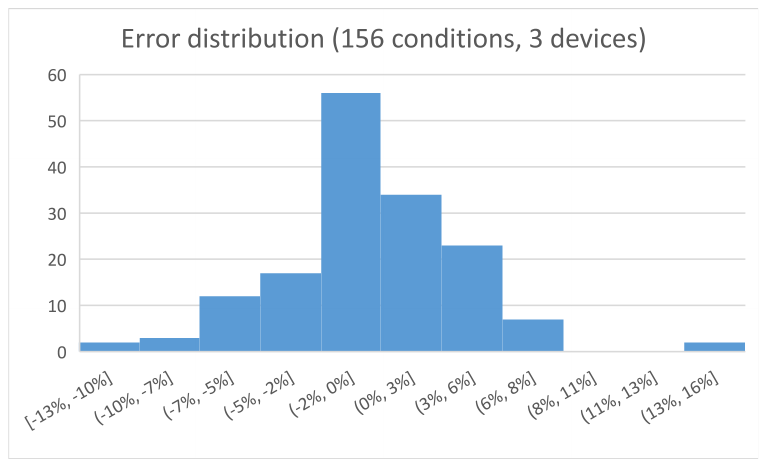

Average: 0%, Standard deviation: 4%

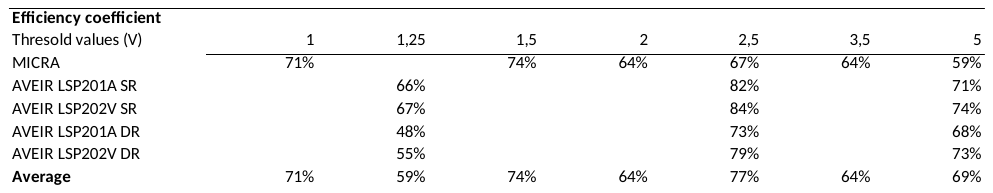

**Cardiac Resynchronisation Therapy - Pacemakers**

| **Modeled longevity vs. nominal longevity provided by manuals** | |
| --- | --- |
| Previous generation 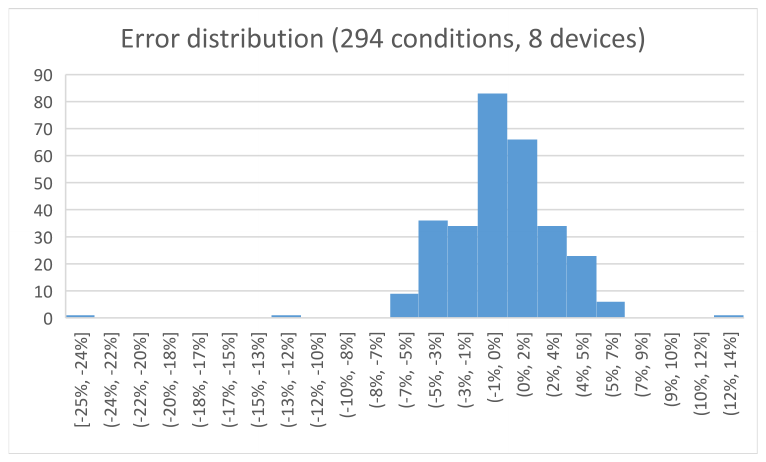 | Commercialized CIEDs 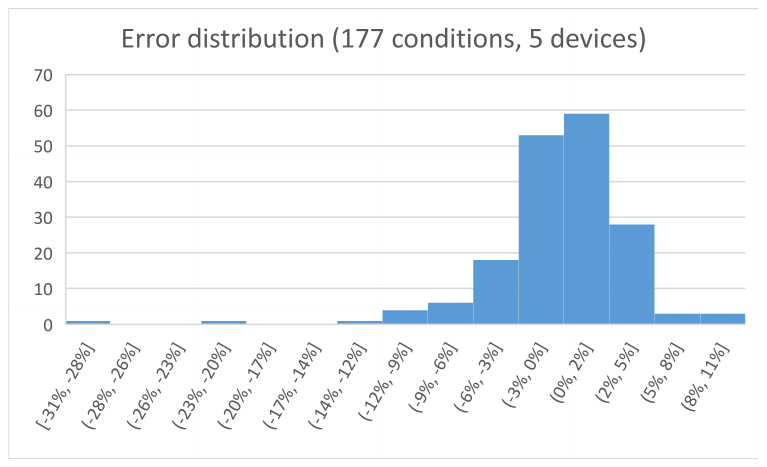 |

| 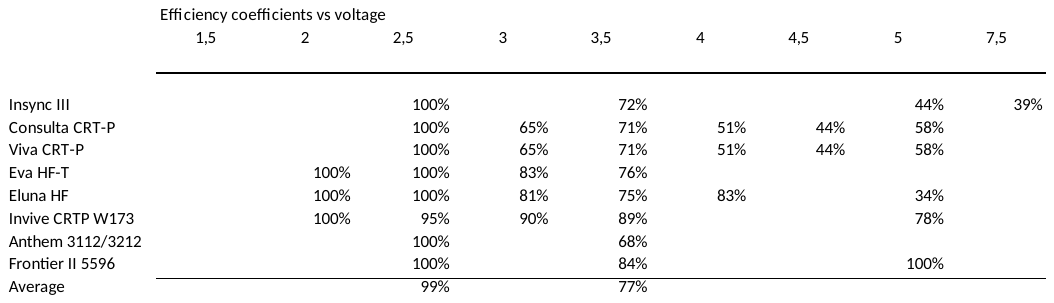 | 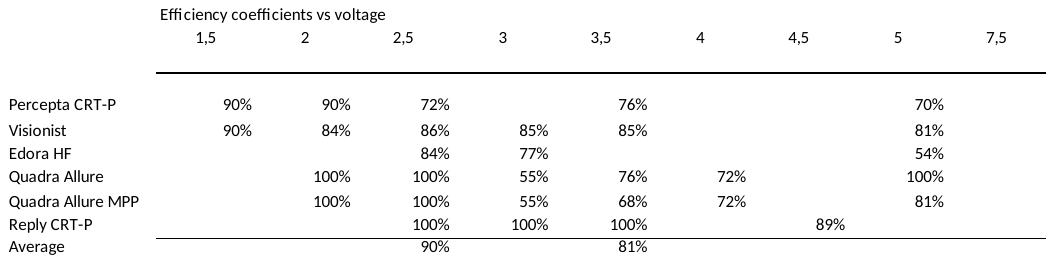 |
| --- | --- |

**3. Pacing current**

The impact of pacing efficiency coefficient on current was evaluated via the current pacing formula:

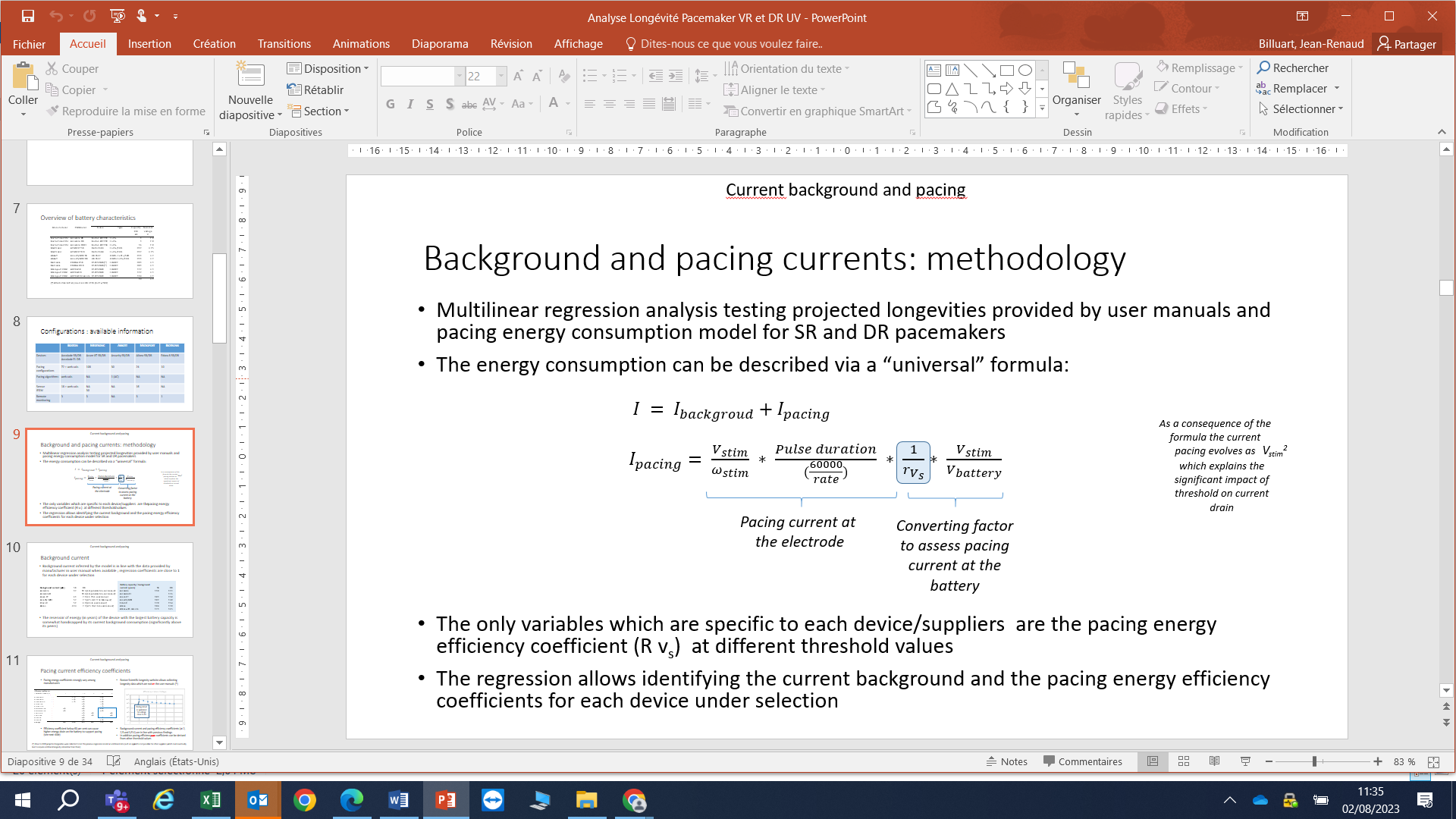

For a given setting (60 bpm, pulse 0.4 ms, 500 ohms, 100% pacing), it is possible to estimate the pacing current depending on pacing efficiency coefficients and battery voltage. The current can be assessed at different battery voltage values (see curves plotted in the chart below). This can be done for each device, according to its own pacing coefficient and battery voltage (see blue dot in the chart below). The dotted red line represents the average current value across devices and the full red lines the interval based on standard deviation values.

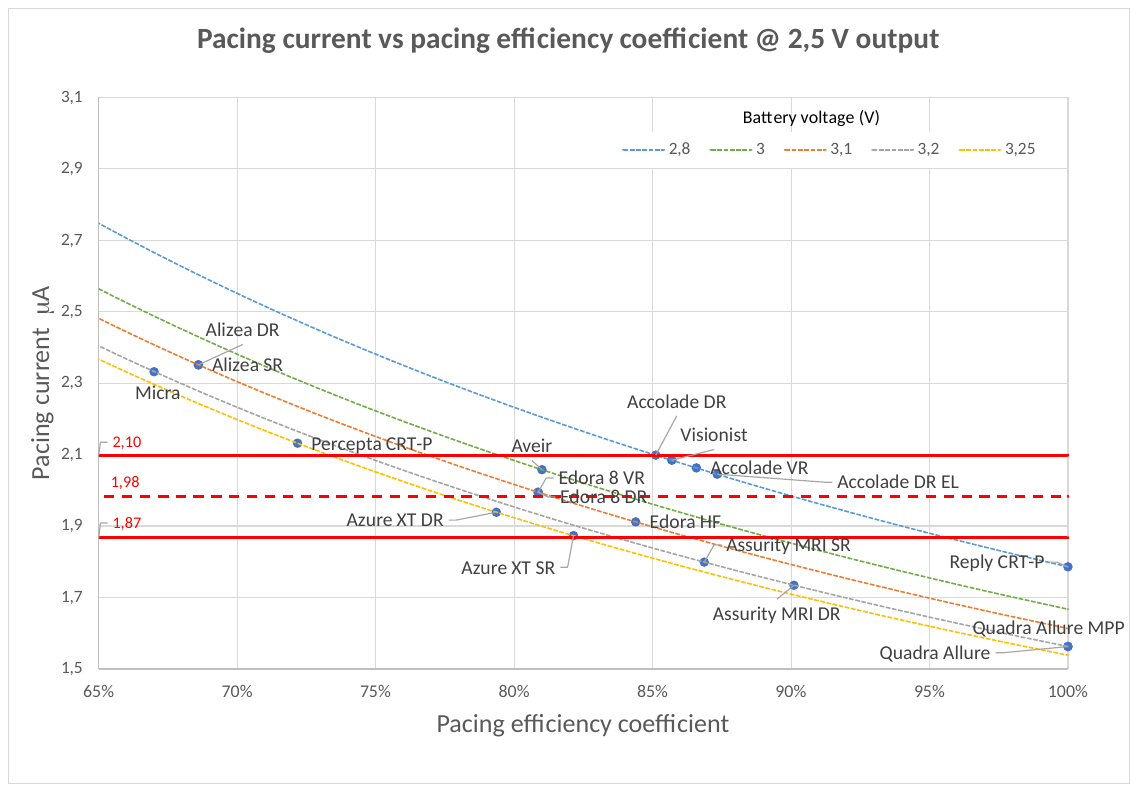

At an output of 2.5V, the average current drain is 1.98+/-0.12μA

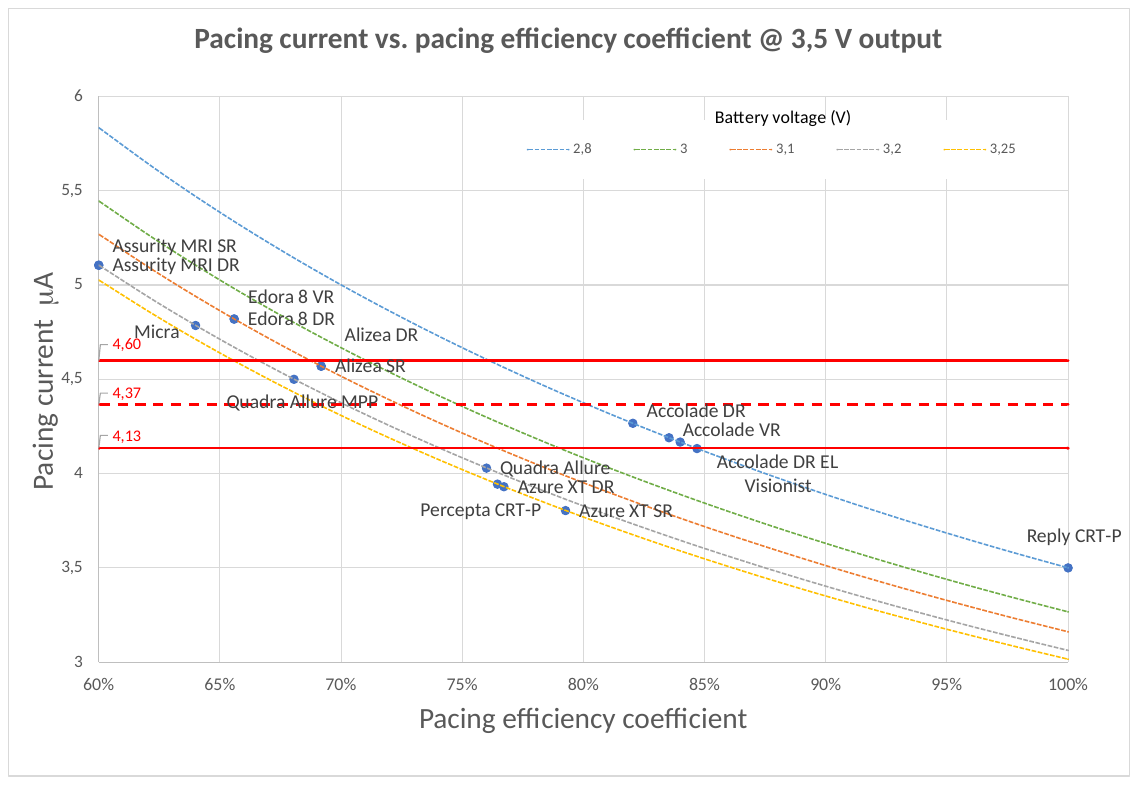

At an output of 3.5V, the average current drain is 4.37+/-0.24μA

**4. Algorithms influencing pacing current**

| **Threshold management** | **Pacing algorithms** |
| --- | --- |
| Threshold alerts (typically used with remote monitoring) which report values but do not automatically adjust outputs relying instead on the manual changes of output are not considered ‘automatic’ algorithms.  Automatic threshold algorithms can be split into two categories: one whose objective is to guarantee capture (with the risk of high pacing outputs), another whose objective is to optimize output (with an optimal target of 1V) while guaranteeing an effective capture.  Within the second group of algorithms, optimization can be done on a daily basis (Capture Management^TM^ from Mdt, Capture Control^TM^ from Btk) or beat to beat (Auto capture^TM^ from Abt, Automatic capture^TM^ from Bsc).  **Manufacturers offering daily optimization do not report energy cost of the algorithm, but the energy cost of the beat to beat optimization algorithm from Abt is available.**  For beat to beat optimization, information from Abt suggests that the energy cost is around **0,82-0,92 μA**. The Bsc longevity calculator was used to derive the impact of this capture algorithm via 72 configurations with the algorithm turned either on or off. Using this methods, the cost of the algorithm from Bsc is close to **1 μA.**  As current background and pacing coefficients were previously established for all voltage for Bsc device, it is possible to conduct a cost benefit analysis between two scenarios (standard pacing at different pacing output vs. an optimal target 1V output with an energy cost of 1 μA reflecting activation of Automatic capture^TM^). The analysis (see below) shows that the Bsc algorithm saves energy (current drain) only if the percentage of pacing is high or the output exceeds 2,5V. Otherwise the algorithm adversely affects (increases) energy consumption.  Capture™ management, available in Micra leadless VVI, is active by default and may contribute to the low capture outputs reported in clinical trials. The energy cost of the algorithm is not reported in the user manual. Auto Capture^TM^ is not available in Aveir™ (possibly because its 1 μA energy cost is too important for a small capsule) | Reduction of Ventricular Pacing (RVP) algorithms have been embedded in all dual chamber pacemaker for 20 years.  Apart from hysteresis mode, each supplier has developed its own algorithm (MVP^TM^ for Mdt, SafeR^TM^ for Mcp, VP suppression^TM^ from Btk, Rythmiq from Bsc), but only two of them (Mdt, Mcp) report an impact on longevity in their user manuals. **The RVP algorithm is reported to have no energy cost overall, saving energy as a result of the reduction of pacing**. The percentage of RVP is not accessible via user manual and can only be identified via clinical studies. A literature review, Aurrichio & al.* concluded that differences existed between algorithms (mainly AAI/DDD vs others) and that AAI/DDD algorithms (MVP^TM^, SafeR^TM^) reduced VP for both SND and AVB. Therefore, even if ventricular pacing algorithms are mainly used in patients with SND (DDD mode is still preferred for AVB) it is possible to assess the potential energy savings for both indications (see table below).  RVP for Aveir DR™ is possible via AV hysteresis.  For recent CRT-P devices, two pacing algorithms impact device energy consumption: Multiple Point Pacing (MMP^TM^) is now available in almost all contemporary devices and a version of Adaptive CRT, (a routinely offered proprietary algorithm from Mdt) is now also commonly available.  For Adaptive CRT™, user manuals report a 50% reduction of RVP for with no energy cost and hence a **reduction of current drain for ventricular pacing by 25%** (from 2x100% for BIV pacing down to 100%+50% for Adaptive CRT pacing).  For MPP, the Abt user manual longevity calculation assumes dual LV pacing. Consequently, **ventricular current pacing increases by 50%** (from 2x100% for BIV to 3x100% for MPP pacing) |

*A. Auricchio & K. A. Ellenbogen, Reducing Ventricular Pacing Frequency in Patients with Atrioventricular Block: is It Time to Change the Current Pacing Paradigm? Circulation: Arrhythmia and Electrophysiology. 2016;9:e004404

**Threshold management (detailed calculations)**

| **Energy used by Autocapture ^TM^ (Abt)** | |
| --- | --- |
| The Assurity DR™ user manual does not describe a significant longevity improvement with Autocapture^TM^ at 1V as compared with standard ventricular pacing at 2.5V. It is possible to derive current consumption related to the Autocapture ^TM^ algorithm (**0.8 – 0.9 µA)** as the difference of energy between these twosettings. | 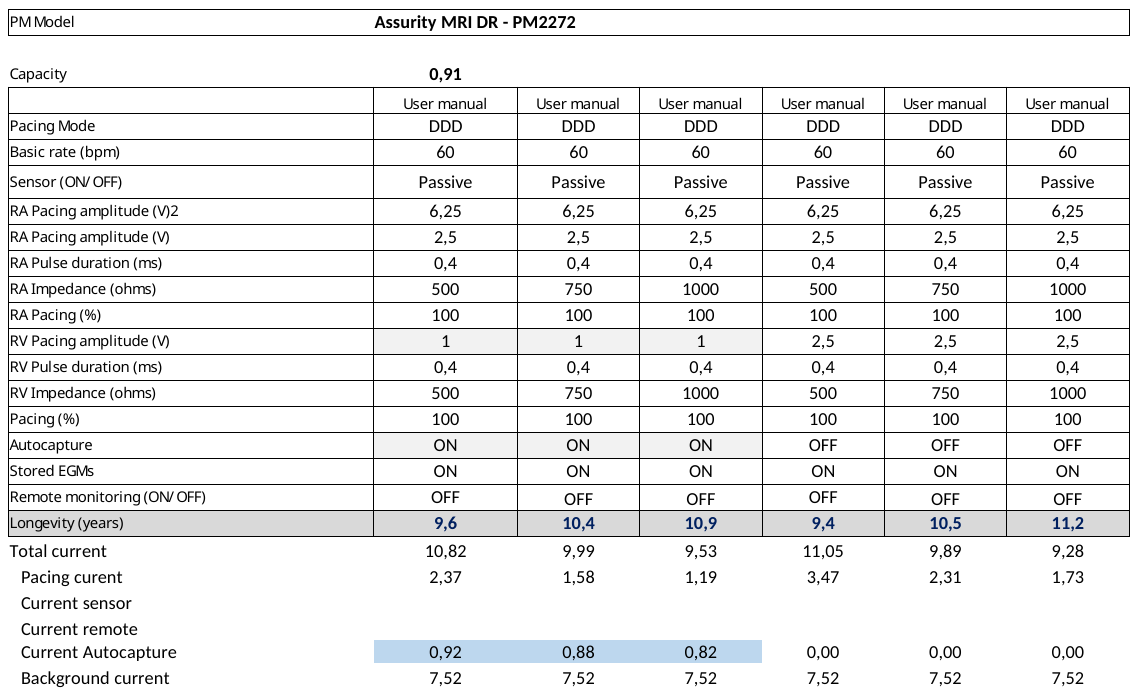 |

| **Energy used by Automatic capture^TM^ (Bsc)** | |
| --- | --- |
| The 72 configurations available in the user manual were tested with the algorithm either on or off. Projected longevities were collected from Bsc longevity calculator website. The difference of longevity offered the opportunity to estimate the current drain for the algorithm (close to 1 µA) | 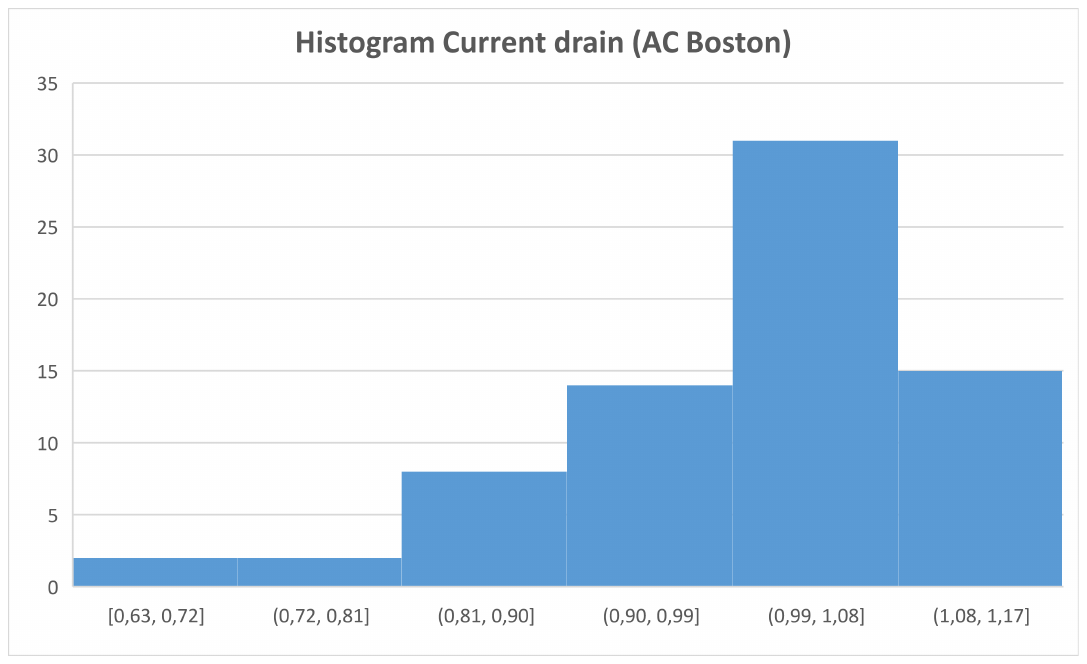 |
| **Energy savings provided by Automatic capture^TM^ (Bsc)** The analysis is conducted for Accolade pacemaker whose pacing coefficients could be estimated for all voltage | |
| *Current drain (μA) of the Accolade™ one chamber device with the algorithm deactivated* | 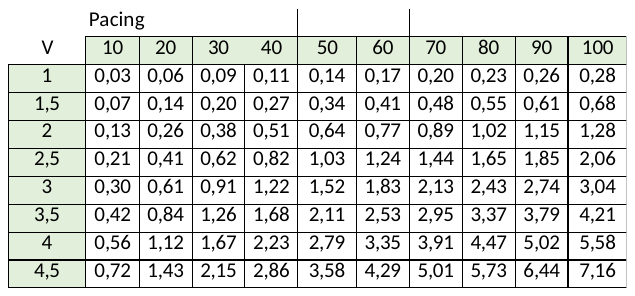 Basic rate 60 bpm, Pulse duration 0.4 ms, 500 ohms |
| *Current drain (μA) of the Accolade™ one chamber device with the algorithm activated* | 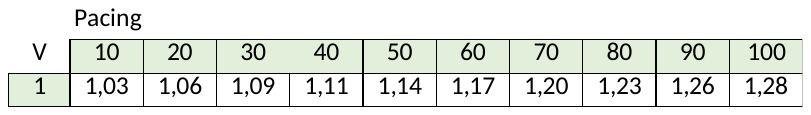 Basic rate 60 bpm, Pulse duration 0.4 ms, 500 ohms, optimal ouptput = 1 V, additional current 1 µA for activation of the algorithm |
| *Difference of current drain algorithm on vs. off*  Values in green highlight savings in terms of energy. Negative values show situations where reduction of current drain with the algorithm does not compensate for the fixed 1 µA energy cost of the algorithm. The more pacing is required and the higher the threshold, the greater the saving. | 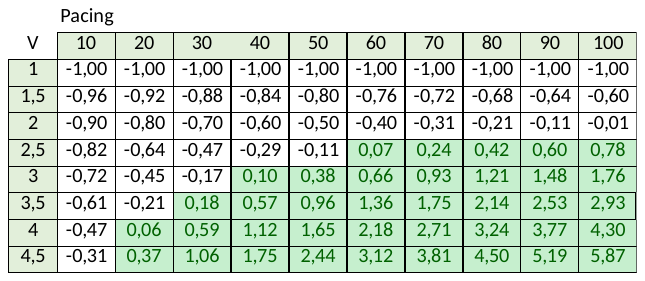 |

**Pacing algorithms (detailed calculations)**

|  | **RVP algorithm (DDD(R))** | | **CRT algorithms** | |
| --- | --- | --- | --- | --- |
| **% of pacing** | **AAI-DDD mode** | **Other RVP mode** | **Adaptive CRT** | **MPP pacing** |
|  | ***Right ventricle*** | | | ***Left ventricle*** |
| **200%** |  |  |  | All pts |
| **100%** | Complete AVB pts | Complete AVB pts |  |  |
| **75%** |  | Inter AVB pts | pts PR>230ms |  |
| **45%** | Intermediate AVB pts |  |  |  |
| **30%** |  |  |  |  |
| **5%** | SND pts | SND pts | pts PR<230ms |  |
| **Average** | **30%** | **45%** | **45%** | **200%** |

(*) With MPP pacing, LV lead delivers 2 points of pacing, AVB; atrioventricular block, SND; sinus node disease.

| **I_pacing_ depending on % of pacing** | **Settings : 60 bpm, pulse 0.4 ms, 500 ohms** | |
| --- | --- | --- |
|  | **Output 2,5 V** | **Output 3,5 V** |
| **200%** | **3,96 μA** | **8,74μA** |
| **100% (†)** | **1,98μA** | **4,37μA** |
| **75%** | **1,49μA** | **3,28μA** |
| **45%** | **0,89μA** | **1,97μA** |
| **30%** | **0,59μA** | **1,31μA** |
| **5%** | **0,10μA** | **0,22μA** |

(**†**) See I_pacing_ values in previous *3. Pacing current*

**5. Other optional features**

The current I is the sum (I = I_background_ + I_pacing_ +I_remote_ + I_IEGM_ + I_sensor_) of all the currents used to maintain the device (I_background_), provided pacing (I_pacing_) and support optional features (remote monitoring and downloads, IEGM, sensor). Currents I_remote_, I_IEGM_, I_sensor_ were estimated via the difference of total current I for similar pacing parameters (I_pacing_ remained unchanged) while corresponding parameters are either turned on or off.

| **IEGM storage** | **Sensor** | **Remote monitoring** |
| --- | --- | --- |
| For most suppliers, IEGM storage is embedded as a standard functionality and energy cost related to IEGM is included already in the current background.  **Only Mdt reports a specific impact on longevity.**  Two options are considered. One option is pre-arrhythmia EGM storage for a period of 6 months (2 x3-month follow-up intervals 6-month period over the life of the device) and a second option is the additional use of Pre-arrhythmia EGM storage (without any period specified).  User manuals indicate that longevity calculations consider the 6 month period as a standard and calculated current drain remains minimal (0.20 – 0.34μA for Enrythm™, 0.03 – 0.20μA for Azure™). Additional use of pre-arrhythmia EGM reduces projected service life by approximately by 34% or 4 months per year for Enrythm™ (equivalent to 5.7-6.53 μA) and by approximately 12.1%-13.8% (equivalent to 0.83- 2.04 μA) for Azure.  For CRT-P, standard IEGM storage costs 0.1-0.48 μA and additional IEGM decreases longevity by 22% (equivalent to 2.6-5.09 μA).  Micra™ leadless memory allows limited IEGM storage (up to 24 hours) and Abt does not report the energy cost of EGM storage in its leadless device. | Rate adaptive pacing usually relies on the G-sensor (accelerometer) to adapt pacing rate according to effort.  For some suppliers, adaption of pacing rate can be optionally enhanced with the combination of a Minute Ventilation sensor (MV-sensor). The longevity calculation provided by user manuals with the MV sensor on or off suggests a fixed energy cost of **0.69 μA** for Bsc and **0.77 μA** for Mcp. Other suppliers do not provide specific data related to sensors. The energy drain for the sensor is similar for CRT-P implants.  Leadless pacemakers Micra™ have only the G sensor.  Activation of sensor leads to an increase of pacing depending on patient’s activities which has not been thoroughly investigated in literature and, to our knowledge is unpredictable. | Remote monitoring did not exist on previous generations devices. For new generation devices, all suppliers, except Abt, provide an estimate of longevity reduction depending on the frequency of transmission. A linear model) is used to derive the fixed cost of remote monitoring (related to the difference between face-to-face device checks and those delivered by the home monitor) and the variable cost (related to the number of transmissions).  The chart below compares the remote monitoring according to manufacturer.  For 2-4 transmission per year, the current consumption is around **1.14-1.75 μA** for RF solutions (BsC, Btk) while it is **0.09-0.59 μA** for the Bluetooth solution. Btk provides a unique solution as its devices transmit on a daily basis (alerts are managed via its website) with a fixed energy cost close to 1.75 μA.  Currently conventional remote monitoring is not available for leadless pacemaker. **Inductive telemetry is possible via radiofrequency (RF)* but Mdt specifies that enabling holter telemetry results in a higher (unspecified) consumption of Micra battery device.** |

**(*) https://www.youtube.com/watch?v=zG28b-EtLAw**

**IEGM storage (source: Mdt user manuals)**

|  | **IEGM storage** | **Extended IEGM storage** |
| --- | --- | --- |
| **SR/DR** |  |  |
| Adapta™ SR/DR | - NA | - NA |
| Enpulse DR™ | - NA | - NA |
| Enrythm DR™ | - IEGM 6 months (2x3m) - Estimated impact = 0.24 μA (0.20-0.35) | - Additional IEGM reduces service life by 34% (4 m per y. - Estimated impact 5.93 µA (5.7-6.53) |
| Azure XT SR/DR™ | - IEGM 6 months (2x3m) - Estimated impact = 0.06 μA (0.03-0.20) | - Additional IEGM decreases longevity by 12-14% (1.4-1.6 m per y) - Estimated impact = 1.10 μA (0.83-2.04) |
| **CRT-P** |  |  |
| Consulta™ | - IEGM 6 months (2x3m) - Estimated impact = 0.27 μA (0.1-0.48) | - Additional IEGM decreases longevity by 22% (2,6 m per y) - Estimated impact = 3.51 μA (2.6-5.09) |
| Viva™ | - IEGM 5 months - Estimated impact = 0.26 μA (0.12-0.5) | - Additional IEGM (18m) decreases longevity by 22% (2.6 m per year) - Estimated additional impact = 3.46 μA (2.7-4.6) |
| Percepta™ | - IEGM off increase longevity by 3,2% or 12 days per year - Estimated impact = 0.43 μA (0.31-1.22) | - NA |

Methodology: if longevity is reduced by 20% by activation of IEGM, according to the longevity formula (L=C/I) current must be increased by 1/(1-20%)). The additional current needed for additional IEGM is the difference I/(1-20%) – I.

**Sensors (source: user manuals)**

Accolade™ 0.69 μA +/- 0,05

Alizea™ 0.78 μA +/- 0.02

**Remote monitoring and downloads**

In user manuals, manufacturers declare longevity for different settings of remote programming which usually include several transmissions per year. We estimated the current drain at different settings. A linear model was used to derive the fixed energy cost (between face-to-face and home monitor and daily checks) and the variable energy cost (related to transmissions). The tables below show the analysis and the result per manufacturer.

| **Boston Scientific LATITUDE^TM^ (RF solution)** | |
| --- | --- |
| 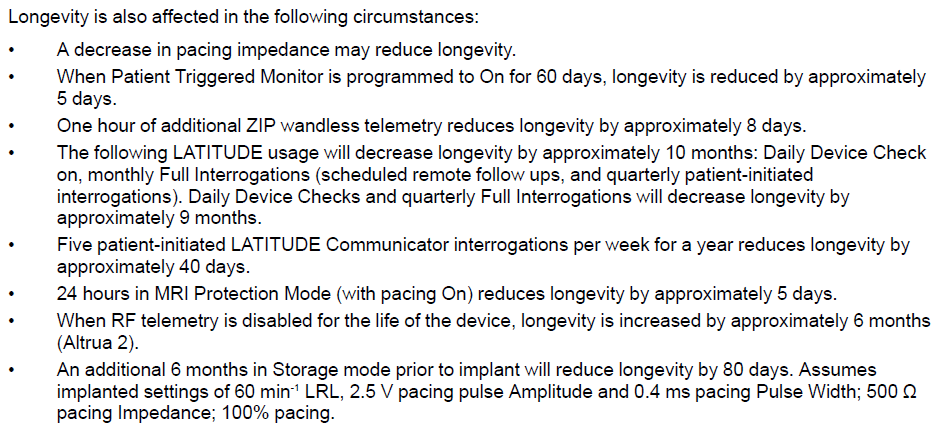 | 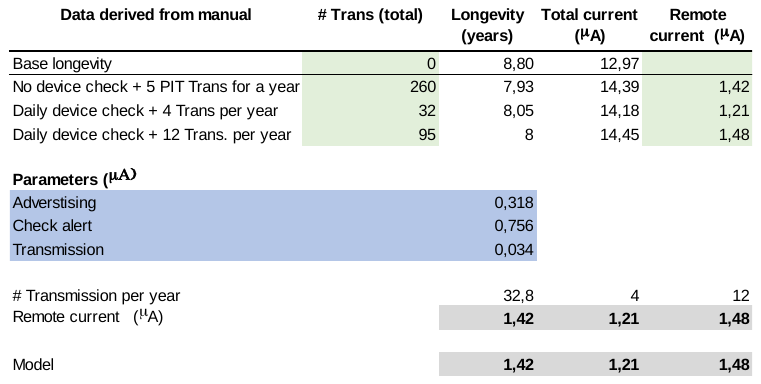 |
| **Medtronic My CareLink ^TM^ (BLE solution)** | |
| *MANUALS*  *Medtronic remote monitor transmissions*  *Additional remote monitoring transmissions reduce the projected service life of the device.*  *For example, from nominal pacing (at 2.5 V, 0.4 ms, 600 Ω, 60 min–1, 100% atrial and ventricular pacing), a patient can expect*  *a projected service life of 10.7 years.*  *More frequent remote monitoring transmissions will reduce this projected service life as follows:*  *Monthly transmissions over the life of the device reduce service life by 16 days, or <1%.*  *Weekly transmissions over the life of the device reduce service life by 87 days, or 2.2%.*  *Daily transmissions over the life of the device reduce service life by 564 days, or 14.4%.* | 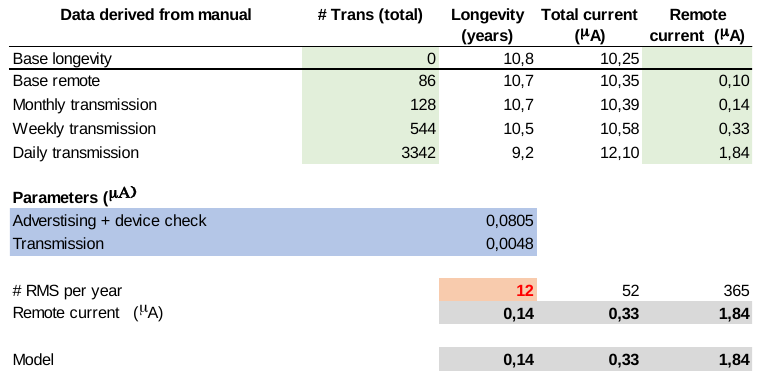 |
| **Biotronik Home Monitoring ^TM^ (RF solution)** | |
| *MANUALS*  *•Home Monitoring ON reduces the service time by approximately 15% in single- and dual-chamber devices and*  *by approximately 15% in single- and dual-chamber devices and by approximately 10% in triple-chamber devices.* | 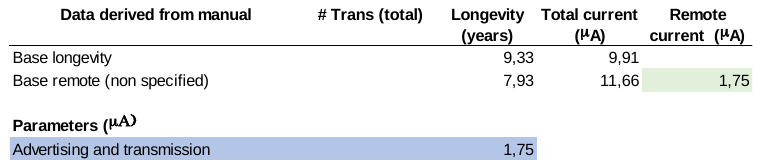 |
| **Microport Smartview ^TM^ (BLE solution)** | |
| *MANUALS*  *• Smartview^TM^ reduces the service time by approximatively:*  *6% for quarterly interrogation*  *21% for weeklu interrogation*  *starting from a service time of 13,7 years (Alizea with remote activated but no transmission)* | 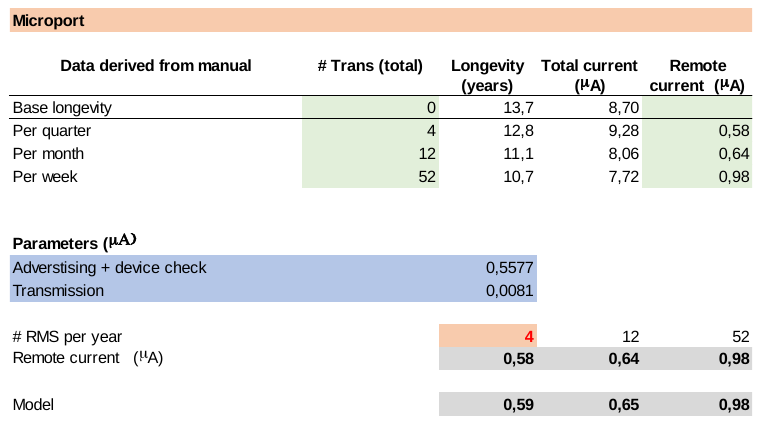 |

Note: Abbott’s user manuals do not report impact of remote monitoring on longevity (we have assumed that it is similar to other RF alternatives such as that of Bsc)

**Comparison between manufacturers**

*MANUALS*

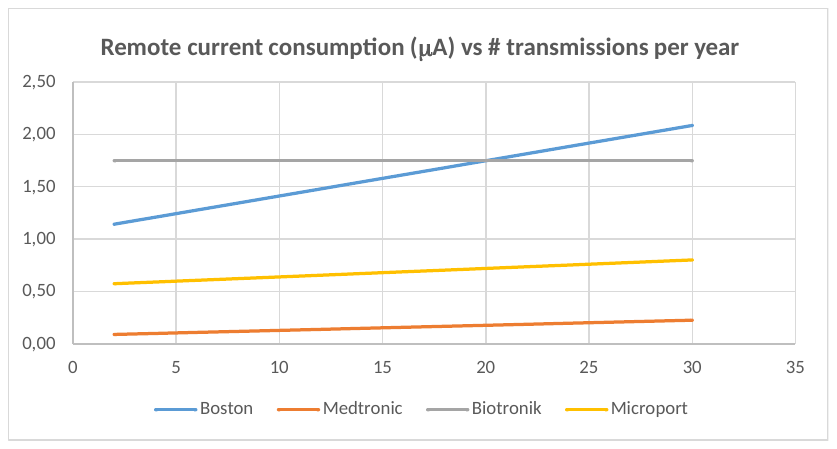

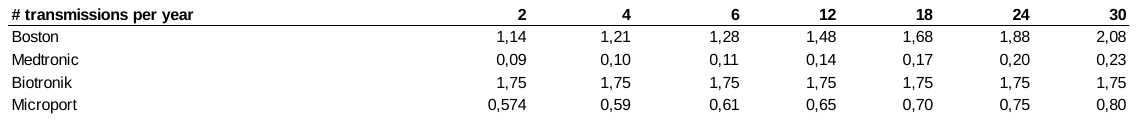

**Leadless pacemakers**

| **Medtronic My CareLink Smart Patient Monitor ^TM^ (Inductive RF communication)** | |
| --- | --- |
| *Inductive telemetry communication is used for the communication between the device and a Medtronic patient connector, which communicates back to a Medtronic clinician device manager to interrogate and program the device. All information is protected in transit by security controls, which include following best practices in regards to proximity and physical security*  *Holter Telemetry – This function allows the implanted device to transmit an EGM with marker telemetry continuously for up to 24 hours, regardless of the use of the patient connector. Enabling Holter Telemetry results in a higher consumption of the device battery.Use of a customized Holter monitor (provided by Medtronic) is required for monitoring the EGM.* | *The impact of inductive RF communication is not specified. It is probably close to 1-2* *μA for RF which is substantial considering Micra has a lower battery capacity when compared with conventional pacemakers* |
