## Supplementary material for "Cardiac Implantable Electronic Devices’ longevity: A novel modelling tool for estimation and comparison": Tables (from another version of this preprint: version 1, 2025-03-31)

**Table 1:** Cardiac Implantable Electronic Device models

|  | **Previous generations:** | **Current CIEDs:** | **Leadless CIEDs** |
| --- | --- | --- | --- |
| *SR* | Abbott (Identity SR™ Adx Model 5180), Biotronik (Evia SR™), Boston Scientific (INSIGNIA I ULTRA™ - 1190), Medtronic (Adapta SR™, ENPULSE™), Microport CRM (Symphonie SR™), and Vitatron (GR20 SR™). | Abbott (Assurity MRI VR™), Biotronik (Edora 8 VR™), Boston Scientific (Accolade VR™), Abbott (Frontier II 5596, Anthem™ 3112/3212), Medtronic (Azure XT VR™), Microport CRM (Alizea VR™ and Alizea VR remote™) | Abbott (Aveir V™ 1.5V and 2.5V), Medtronic (Micra™ 1.5V and 2.5V) |
| *DDD* | Abbott (Identity XL DR™ Model 5386 and Identity DR™ Model 5380), Biotronik (Evia DR™ and Evia DR-T™), Boston Scientific (INSIGNIA I ULTRA™ - 1291), Medtronic (ENPULSE E2DR™ 31/33, Enrythm DR™, Adapta DR™, ENPULSE E2 DR21™), Microport CRM (Symphonie DR™), and Vitatron (GR70 DR™) | Abbott (Assurity MRI DR™), Biotronik (Edora 8 DR™), Boston Scientific (Accolade DR EL™ and Accolade DR™), Medtronic (Azure XT DR™), Microport CRM (Alizea DR™ and Alizea DR remote™) | Abbott (Aveir V™ 1.5V and 2.5V, Aveir A™ 1.5V and 2.5V) |
| *CRT-P* | Abbott (Frontier II 5596™, Anthem 3112/3212™), Biotronik (Eva HF-T™, Eluna HF™), Boston Scientific (Invive CRT-P W173™), Medtronic (Consulta CRT-P™, Viva CRT-P™, InsyncIII™) | Abbott (Quadra Allure™, Quadra Allure MPP™), Biotronik (Edora HF™, Eluna HF™), Boston Scientific (Visionist™), Medtronic (Percepta CRT-P aCRT™, Percepta CRT-P ™), Microport CRM (Reply CRT-P™) |  |

**Table 2:** Nominal voltage and capacity by Cardiac Implantable Electronic Device model, according to manufacturers’ manuals

|  |  | Previous generation | Nominal voltage (V) | Capacity  to ERI  (Ah) |  |  | Current generation | Nominal voltage (V) |  | Capacity  to ERI  (Ah) |
| --- | --- | --- | --- | --- | --- | --- | --- | --- | --- | --- |
| **SR** | Abt | Identity™ SR Adx Model 5180 | 2.80 | 0.55 |  | Abt | Assurity MRI SR™ | 3.20 |  | 0.91 |
|  | Btk | Evia™ SR | 2.80 | 1.20 |  | Btk | Edora 8 SR™ | 3.10 |  | 0.81 |
|  | BSc | INSIGNIA™ I ULTRA-1190 | 2.80 | 0.97 |  | BSc | Accolade VR™ | 2.80 |  | 1.00 |
|  | Mcp | Symphonie™ SR | 2.80 | 0.93 |  | Mcp | Alizea SR™ | 3.10 |  | 1.12 |
|  | Mdt | Enpulse ™ E2SR/Adapta SR | 2.80 | 0.86/0.86 |  | Mdt | Azure XT SR™ | 3.25 |  | 0.97 |
|  | Vit | G20 SR | 2.80 | 0.86 |  |  |  |  |  |  |
| **DR** | Abt | Identity™ D 5380/DR | 2.80 | 0.55/0.95 |  | Abt | Assurity MRI DR™ | 3.20 |  | 0.91 |
|  | Btk | Evia™ DR 2 models DR-T/DR | 3.1/2.8 | 1.05/1.2 |  | Btk | Edora 8 DR™ | 3.10 |  | 0.81 |
|  | BSc | Insignia™ ULTRA 1290/1291 | 2.80 | 0.935/1.44 |  | BSc | Accolade™ DR, DR-EL | 2.80 |  | 1/1.6 |
|  | Mcp | Symphonie™ DR | 2.80 | 0.93 |  | Mcp | Alizea DR™ rem/no rem | 3.10 |  | 1.04/1.12 |
|  | Mdt | Enpulse™21/Adapta™/Enpulse™33 | 2.80 | 0.82/1.2/1.4 |  | Mdt | Azu XT DR™ | 3.25 |  | 0.97 |
|  | Mdt | Enrythm™ DR | 3,20 | 1.10 |  |  |  |  |  |  |
|  | Vit | G70™ DR | 2.80 | 1.22 |  |  |  |  |  |  |
| **CRT-P** | Abt | Anthem™ 3112-3212/Frontier™ II | 3.2/2.8 | 0.8/0.95 |  | Abt | Quadra Allure™ | 3.2 |  | 0.8 |
|  | Btk | Evia™ HF-T/Eluna™ HF | 3.1 | 1.0 |  | Btk | Edora HF™ | 3.1 |  | 0.932 |
|  | BSc | Invive™ CRTP W173 | 3 | 1.36 |  | BSc | Visionist™ | 2.8 |  | 1.5 |
|  | Mdt | Insync III™ | 3.25 | 1.4 |  | Mcp | Reply CRT-P™ | 2.8 |  | 0.82 |
|  | Mdt | Consulta CRT-P™/Viva CRT-P™ | 3.2 | 0.97 |  | Mdt | Percepta CRT-P™ | 3.25 |  | 1.03 |

Note: Btk is the only supplier pursuing multiple battery sources, but according to the Btk user manuals, differences in battery capacity have no impact on device longevity.

|  |  | Leadless pacemakers | Nominal voltage (V) | Capacity ERI  (Ah) |
| --- | --- | --- | --- | --- |
|  | Abt | AVEIR LSP201™ Atrial capsule | 3.0 | 0.174 |
|  | Abt | AVEIR LSP202™ Ventricular capsule | 3.0 | 0.241 |
|  | Mdt | MICRA™ | 3.2 | 0.12 |

**Table 3:** Background currents (from manufacturer’s manuals and modelling) by device

|  |  | | Previous generation CIED models | | | Reported background current (μA) according to manuals | | Modeled background current  (μA) | |  |  | | Current CIED models | | | Reported background current (μA) according to manuals | | | Modeled background current  (μA) |
| --- | --- | --- | --- | --- | --- | --- | --- | --- | --- | --- | --- | --- | --- | --- | --- | --- | --- | --- | --- |
| **SR** | Abt | | Identity™ SR Adx Model 5180 | | | 6.30 | | 5.94 | |  | Abt | | Assurity™ MRI SR | | | 5.40 | | | 5.70 |
|  | Btk | | Evia™ SR | | | 6.00 | | 7.21 | |  | Btk | | Edora™ 8 SR | | | 6.00 | | | 5.20 |
|  | BSc | | INSIGNIA I ULTRA - 1190 | | | NA | | 10.72 | |  | BSc | | Accolade™ VR | | | NA | | | 9.70 |
|  | Mcp | | Symphonie™ SR | | | NA | | 5.78 | |  | Mcp | | Alizea™ SR | | | 5.73 | | | 5.80 |
|  | Mdt | | Enpulse ™ E2SR/ Adapta SR | | | 11/12.93 | | 11.1/11.34 | |  | Mdt | | Azure™ XT SR | | | 6.71 | | | 6.10 |
|  | Vit | | G20™ SR | | | 9.80 | | 9.14 | |  |  | |  | | |  | | |  |
| **DR** | Abt | | Identity™ D 5380 / DR | | | 6.90 | | 6.19/5.72 | |  | Abt | | Assurity™ MRI DR | | | 7.70 | | | 7.50 |
|  | Btk | | Evia™ DR 2 models DR-T / DR | | | 6.00 | | 6.66/9.86 | |  | Btk | | Edora™ 8 DR | | | 6.00 | | | 6.00 |
|  | BSc | | Insignia™ ULTRA 1290/ 1291 | | | NA | | 11.62/13.06 | |  | BSc | | Accolade™ DR, DR-EL™ | | | NA | | | 10.3/10.4 |
|  | Mcp | | Symphonie™ DR | | | NA | | 6.00 | |  | Mcp | | Alizea™ DR rem /no rem | | | 6.00 | | | 6.30 |
|  | Mdt | | Enpulse™ 21/ Adapta™/ Enpulse™ 33 | | | 13.3/13.87/13.3 | | 13.28/14.67/13.7 | |  | Mdt | | Azure™ XT DR | | | 6.71 | | | 7.00 |
|  | Mdt | | Enrythm™ DR | | | 9,80 | | 9,80 | |  |  | |  | | |  | | |  |
|  | Vit | | G70™ DR | | | 10,00 | | 11,42 | |  |  | |  | | |  | | |  |
| **CRT-P** | Abt | | Anthem™ 3112-3212 / Frontier II | | | NA | | 7,09 | |  | Abt | | Quadra Allure™ | | | NA | | | 7.09 |
|  | Btk | | Evia™ HF-T/Eluna HF | | | 7.0 | | 7,26/7,56 | |  | Btk | | Edora HF™ | | | 7.0 | | | 7.1 |
|  | BSc | | Invive™ CRTP W173 | | | NA | | 11,77 | |  | BSc | | Visionist™ | | | NA | | | 9.63 |
|  | Mdt | | Insync™ III | | | 12.0 | | 11,90 | |  | Mcp | | Reply CRT-P™ | | | 7.9 | | | 7.5 |
|  | Mdt | | Consulta™ CRT-P/Viva CRT-P | | | 7.07/7.22 | | 7,06/7,22 | |  | Mdt | | Percepta CRT-P™ | | | 7.14 | | | 7.19 |

|  |  | Leadless pacemakers | Background current  (μA) from manuals | Modeled background current  (μA) |
| --- | --- | --- | --- | --- |
| **SR** | Abt | AVEIR™ LSP201Atrial capsule | 0.94 | 1.06 |
|  | Abt | AVEIR™ LSP202Ventr. Capsule™ | 0.94 | 1.02 |
|  | Mdt | MICRA™ | 0.8 | 0.94 |
| **DR** | Abt | AVEIR™ LSP201Atrial capsule | 1.8 | 2.03 |
|  | Abt | AVEIR™ LSP202Ventr. capsule | 1.8 | 1.94 |

**Table 4:** Recommendations on device selection and programming

|  | **Device selection prior implant** | **Programming post implant** |
| --- | --- | --- |
| SR | - Avoid Accolade™ SR | - Use capture algorithm to reduce pacing output |
| DR | - Avoid Accolade™ DR (non-EL version) | - Use AAI/DDD mode not only for SND patients. but also, intermittent AVB patients. - Use capture algorithm to reduce pacing output if % of RVP > 60% |
| CRT-P | - Prefer Visionist™, Percepta™ (+aCRT™) | - Use capture algorithm to reduce pacing outputs (RV capture is inefficient if combined with aCRT™) - Avoid MMP pacing |
| IEGM |  | - Avoid extended IEMG storage |
| Remote monitoring | - Anticipate “one off” energy cost for some devices (Edora™, Alizea™) | - Avoid using inductive telemetry for leadless pacemaker |
| Leadless VVI |  | - Use capture algorithm to reduce pacing output or carefully program pacing outputs if capture algorithm is not available |
| Leadless DR | - Consider intermittent AVB patients and anticipate early replacement of atrial capsule for SND patients. | - Idem (see above) |
